# AI-Driven Predictive Biomarker Discovery with Contrastive Learning to Improve Clinical Trial Outcomes

**DOI:** 10.1101/2024.01.31.24302104

**Authors:** Gustavo Arango-Argoty, Damian E. Bikiel, Gerald J. Sun, Elly Kipkogei, Kaitlin M. Smith, Sebastian Carrasco Pro, Etai Jacob

## Abstract

Modern clinical trials can capture tens of thousands of clinicogenomic measurements per individual. Discovering predictive biomarkers, as opposed to prognostic markers, is challenging when using manual approaches. To address this, we present an automated neural network framework based on contrastive learning—a machine learning approach that involves training a model to distinguish between similar and dissimilar inputs. We have named this framework the Predictive Biomarker Modeling Framework (PBMF). This general-purpose framework explores potential predictive biomarkers in a systematic and unbiased manner, as demonstrated in simulated “ground truth” synthetic scenarios resembling clinical trials, well-established clinical datasets for survival analysis, real-world data, and clinical trials for bladder, kidney, and lung cancer. Applied retrospectively to real clinicogenomic data sets, particularly for the complex task of discovering predictive biomarkers in immunooncology (IO), our algorithm successfully found biomarkers that identify IO-treated individuals who survive longer than those treated with other therapies. In a retrospective analysis, we demonstrated how our framework could have contributed to a phase 3 clinical trial (NCT02008227) by uncovering a predictive biomarker based solely on early study data. Patients identified with this predictive biomarker had a 15% improvement in survival risk, as compared to those of the original trial. This improvement was achieved with a simple, interpretable decision tree generated via PBMF knowledge distillation. Our framework additionally identified potential predictive biomarkers for two other phase 3 clinical trials (NCT01668784, NCT02302807) by utilizing single-arm studies with synthetic control arms and identified predictive biomarkers with at least 10% improvement in survival risk. The PBMF offers a broad, rapid, and robust approach to inform biomarker strategy, providing actionable outcomes for clinical decision-making.

## INTRODUCTION

The promise of precision medicine lies in treating patients with therapies that precisely target their unique diseases.^1,2^ Using biomarkers to select a study population more likely to benefit from a therapeutic effect is crucial for increasing the efficiency of clinical trials in demonstrating effectiveness.^3^ For example, the impact of biomarkers on drug development is highlighted by compelling findings in the BIO 2021 report,^4^ which shows that drug development programs integrating patient preselection biomarkers have a striking two-fold increase in the likelihood of approval, reaching 15.9%. However, discovering predictive biomarkers - characteristics that identify individuals more likely to experience a favorable treatment effect compared to those without such characteristics - is a complex and challenging endeavor. The intricate interplay of genetics and environmental factors, coupled with the complexity of disease biology and treatments, makes the discovery of predictive biomarkers a daunting task. The scarcity of comprehensive data, which is often due to acquisition or technical difficulties, presents challenges to the accurate representation of diverse populations, disease subtypes, and treatment cohorts, further compounding this discovery challenge. Moreover, the presence of numerous prognostic factors often hinders the ability to pinpoint the predictive biomarker within the studied patient population. The advent of next-generation sequencing technologies providing large-scale profiling of gene mutations, transcript expression and protein, have both increased our opportunity to find predictive biomarkers as well as further complicated the task.^5^ Finally, even if a putative biomarker is found, translational applicability must be assessed with independent validation cohorts, adding further complexity and cost.

Nevertheless, there are clinically validated predictive biomarkers for certain targeted therapies, exemplified by the identification of *BCR-ABL* and *EGFR* mutations guiding the use of receptor tyrosine kinase inhibitors in cancer treatment.^6^ Despite these significant achievements, a considerable gap remains in the availability of predictive biomarkers, particularly for therapies that indirectly target the disease, like those for immunooncology (IO), which modulates the immune system rather than the tumor, and therefore lacks an obvious molecular biomarker hypothesis. Although PD-L1 expression,^7^ microsatellite instability,^8^ and tumor mutation burden (TMB)^9^ serve as validated predictive biomarkers for IO, only a subset of responsive patients exhibit positivity for these markers.^10^ With an expanding array of novel targeted therapies, immunotherapies, and their combinations under investigation in clinical trials, the development of methodologies for identifying predictive biomarkers becomes imperative to advance precision medicine and optimize the efficacy of emerging treatments.

To address the challenge of predictive biomarker discovery, traditional regression methods such as Cox proportional hazards (PH) modeling^11^ have been widely employed. However, these methods necessitate the explicit enumeration of covariates and interactions, a task that becomes impractical as the number of features increases, particularly in scenarios involving a diverse set of clinical and -omic features. More recently, algorithms have been developed that aim to discover predictive biomarkers without requiring such explicit specifications. These approaches utilize algorithms designed to maximize the difference in target outcomes between subgroups with different treatments.^12,13^ Unfortunately, even these advanced approaches encounter challenges in identifying a predictive signal in the presence of noisy data or features that uniformly influence all arms (i.e., are prognostic) and often result in overfitting.

We therefore developed a novel approach, the predictive biomarker modeling framework (PBMF), designed for end-to-end predictive biomarker discovery and evaluation (Fig. 1). This framework, now available to the research community, centers around a neural network ensemble model featuring a contrastive loss function that ensures the learning of a multivariate biomarker that is specific to a target treatment of interest but not to a control treatment (see description in the results section below). The biomarker score cutoff and sample prevalence constraints are also components of the model’s training objective (loss function), abrogating the need for post-hoc tuning. Additionally, we provide tools for generating simulated data to benchmark the model, along with features to distill the model into an interpretable, deployable biomarker.

**Fig. 1:**
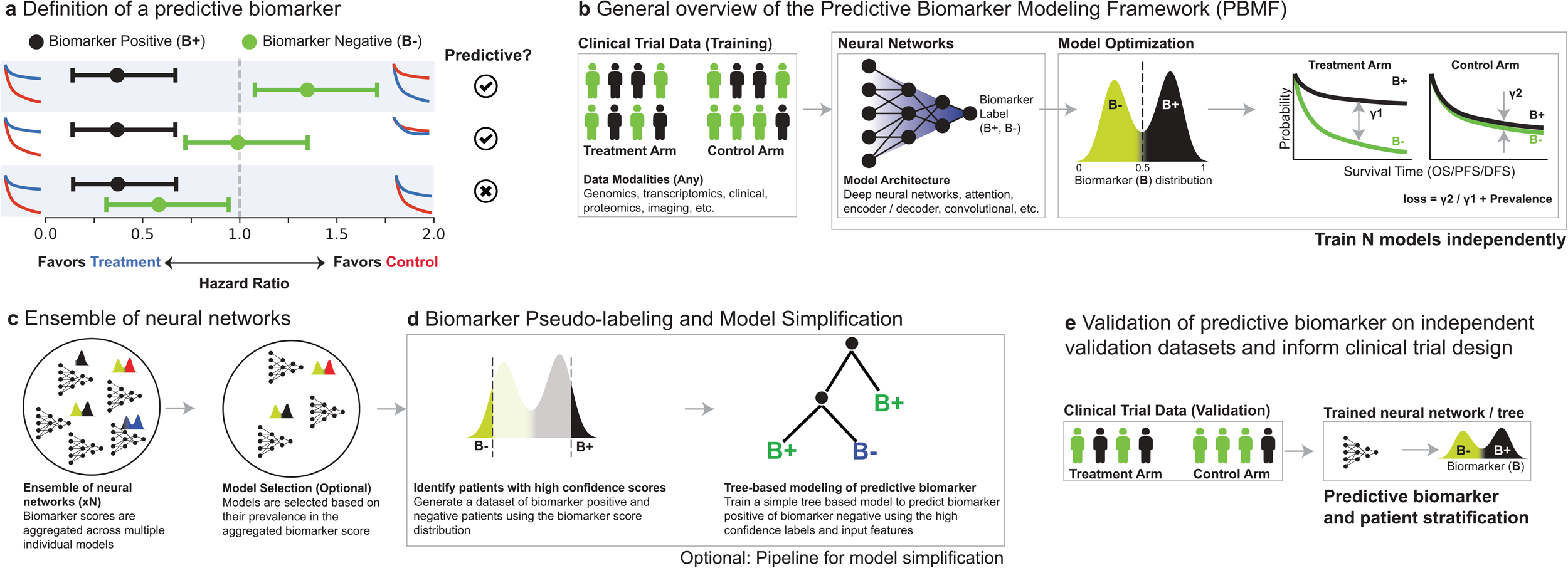
Detailed schematic of the PBMF. **a**, Discrimination between predictive and prognostic biomarkers, with the subdivision into B+ and B− cohorts. B+ is indicative of patients who benefit from treatment as opposed to the control, and B− signifies a lack of superiority of treatment or an advantage in the control group. **b**, The PBMF trains a set (*N*) of neural networks, each independently trained on clinical trial data with a contrastive loss function. The loss is designed to enhance the differential impact of B+ versus B− in the treatment group and concurrently minimize B+ influence over B− in the control arm. **c**, The ensemble of PBMF models synthesizes into a consolidated predictive score, refining the model collection by filtering out non-contributory models to retain only those with significant impact. **d**, High-confidence patient samples are identified through biomarker pseudo-labeling, which then serve to construct an interpretable, simplified decision tree model, categorizing patients as B+ or B−. **e**, External dataset validation of the PBMF model affirms the biomarker’s predictive capacity, demonstrating the model’s reliability from ensemble to simplified tree representation, thus reinforcing its utility in clinical trial stratification. OS, overall survival; PFS, progression-free survival; DFS, disease-free survival.

Here, we provide a diverse body of empirical evidence showcasing the robust predictive biomarker discovery capability of the PBMF across various scenarios, including simulated biomarker discovery, well-established clinical datasets for survival analysis, real-world data, and randomized controlled clinical trials. Notably, the PBMF outperformed existing approaches in subgroup identification within both simulated and real data sets. Furthermore, we illustrate how the PBMF retrospectively contributed to patient selection in a phase 3 clinical trial by uncovering a predictive biomarker based solely on phase 2 trial data. This discovery led to a 15% improvement in efficacy in the original trial, achieved through a straightforward decision tree generated via PBMF knowledge distillation. Finally, we show how the PBMF may also retrospectively contribute to patient selection for two additional phase 3 clinical trials, using only single-arm early phase trial data with synthetic control arms, leading to at least a 10% improvement in efficacy versus the original trials.

## RESULTS

### Predictive biomarkers, contrastive learning, and model architecture

We define a predictive biomarker, B, as a tool categorizing a population into positive (B+) or negative (B−) for the biomarker, specific to a given treatment. B can encompass various patient measurements (e.g., age, blood counts, RNA gene expression). The biomarker is predictive if the B+ subpopulation is selectively enriched for individuals benefitting from a treatment of interest (“treatment”), but not a comparator one (“control”; Fig. 1a). Similarly, the B− subpopulation should be selectively enriched for those not benefiting from any treatment, or perhaps benefiting instead from a comparator (Fig. 1a). In contrast, a prognostic biomarker is characterized by similar benefit irrespective of treatment (Fig. 1a, bottom).

With this definition, we formulated the PBMF to distinguish between two patient populations based on their differential response to treatments, i.e. contrastive learning. Specifically, the training objective of the PBMF (i.e. its loss function) actively maximizes the differences in outcomes for a given treatment (similar to pushing apart dissimilar items in contrastive learning) for B+ versus B− patients. Simultaneously, it minimizes the differences in outcomes for the control arm (similar to bringing similar items closer in contrastive learning). By doing so, the network is trained to contrast the effects of two treatments across the biomarker-defined groups, effectively learning the distinctive features that separate patient responses. More formally from a technical perspective, the loss function is defined as the log difference between control and treatment log-rank test statistics (Fig. 1b; Methods). In plain terms, this has the effect of maximizing the separation of survival curves (or generally, for any time-to-event curves) between B+ and B− in the subpopulation receiving the treatment (i.e. large log-rank test statistic) while minimizing the separation for the subpopulation receiving the control. The model therefore optimizes for predictive biomarker behavior (Fig. 1a, 1b). For applications requiring a particular biomarker prevalence, the PBMF can be run with an optional constraint (specifically, a penalization term) to encourage a predefined B+ prevalence proportion.

We designed the PBMF to be flexible and usable by the technical community (via an application programming interface). In particular, its modular design allows use of any neural network-based machine learning model, including deep, convolutional, and attention-based networks. The PBMF can use data from any modality (e.g., genomics, clinical, imaging), without restriction on the number or type (e.g., categorical or continuous; Fig. 1b). The PBMF outputs a “confidence” (i.e. probability) score from 0 to 1, which can be used (strictly speaking as a likelihood) to assign a sample to the B+ or B− subpopulation.

### Model implementation and extensions

Overfitting poses a significant challenge in biomarker discovery, due to heterogeneity in patient populations and large numbers of features, particularly when attempting to predict the efficacy of one treatment over another rather than that of a single treatment. The PBMF therefore incorporates an established solution to increase model robustness by allowing training of a diverse collection of models (i.e. *n* independently trained neural networks), also known as an ensemble (Fig. 1c, left), and then aggregating the ensemble predictions to yield a better prediction than any ensemble constituent. Model diversity is achieved by allowing each model to learn with a unique random subset of samples and features (akin to the machine learning principle of bagging^14^; Table S1). Following model training, we provide a solution whereby one can optionally remove poor performing models in the ensemble, i.e. model pruning, which can further enhance ensemble performance (Fig. 1c, right).

Finally, an opaque neural network in the PBMF-generated biomarker may compromise confidence and hinder applicability in clinical settings. To address this, the PBMF incorporates an optional pipeline for simplifying the model (‘model distillation’) into a parsimonious, interpretable decision tree. This is achieved by training a decision tree classifier on the subset of samples for which the ensemble had the highest confidence scores (Fig. 1e). This decision tree thus transforms the candidate predictive biomarker into a simple set of rules, facilitating seamless integration into the design of future clinical studies (Fig. 1d, 1e).

### PBMF identification of predictive biomarkers in diverse simulated biomarker discovery scenarios

To facilitate benchmarking, we generated synthetic data sets representing realistic combinations of features and time-to-event data (i.e., survival), mirroring conditions commonly encountered in real-world scenarios (Fig. 2a). Benchmarking was performed across 100 replicates, with performance reported on held-out test data sets from each replicate. We compared performance only across PBMF and Virtual Twins^15^ (VT) methods, as SIDES^16^ (subgroup identification based on differential effect search) failed to solve the simulated scenarios.

**Fig. 2:**
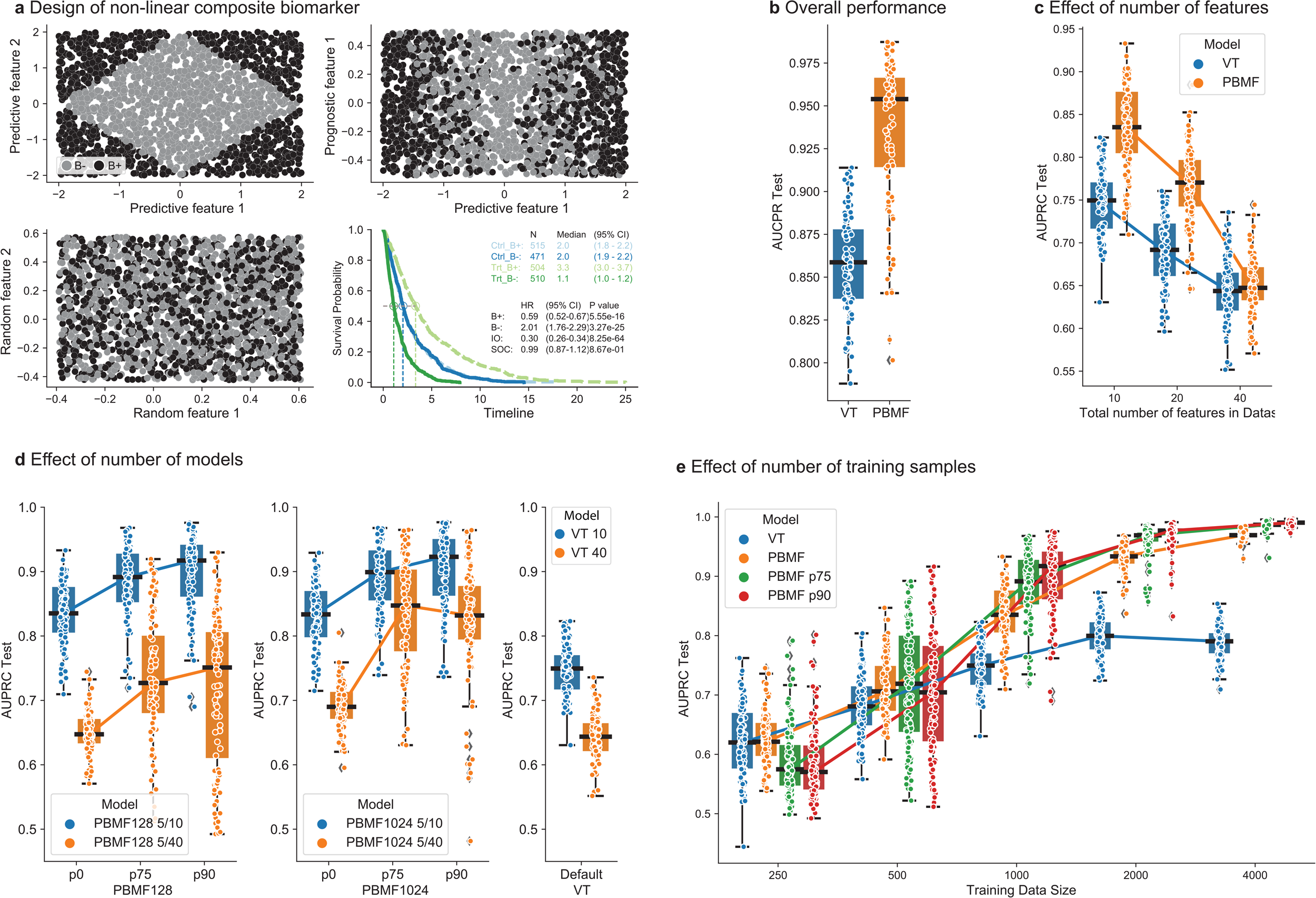
Simulated and benchmark tests. **a**, Synthetic data set generation and behavior. A predictive biomarker is generated by ‘predictive feature 1’ and ‘predictive feature 2’ (top left), which creates a particular Kaplan-Meier plot, showing the differential effect in the treatment (Trt) and control arms (Ctrl; bottom right). ‘Prognostic feature 1’ has a different effect when added to ‘predictive feature 1’ (top right). Random features with no structure can be added (‘random feature 1’ and ‘random feature 2’; bottom left). **b**, AUPRC for the test set comparing the PBMF model developed for a data set containing 3 features (2 predictive, 1 prognostic) in orange against VT in blue. The training was performed on 1000 data points, with 100 training-test split replicates **c**, Effect of the number of random features in the AUPRC for PBMF and VT. The PBMF model contains 128 ensembles of 5 features chosen from data sets with 10, 20, and 40 total features, in which only 2 are predictive and 1 is prognostic. Models are trained with 1000 data points, with 100 training-test split replicates. **d**, Effect of the number of models in the ensemble for PBMF (128 vs 1024) against VT at two different levels of noise (10 and 40 total features; 5 features chosen). Models are trained with 1000 data points, with 100 training-test split replicates. **e**, Effect of the training size on AUPRC for VT (blue), PBMF (orange), and two different levels of post-pruning (top quartile [p75, green] and top decile [p90, red] percentile of models). The data set contained 10 total features (2 predictive, 1 prognostic, and 7 random). PBMF ensemble models comprised 128 models containing only 5 features from the 10. Boxplot: centerline, median; box limits, quartile 1 and 3; box whiskers, 1.5x interquartile range; diamonds, outliers; dots, data points.

The objective of the first benchmarking scenario was to discover a predictive signal in the presence of a prognostic signal. This scenario comprised 3 features, 2 predictive and 1 prognostic; importantly, the predictive signal was present only as a combination of the two predictive features (Fig. 2a). The PBMF yielded an area under the precision-recall curve (AUPRC) of 0.918 ± 0.047 (mean ± standard deviation) and outperformed a competing method, VT (AUPRC = 0.858 ± 0.029) (Fig. 2b, Table S2).

Real-world scenarios often involve the presence of noninformative features, complicating the extraction of the underlying predictive signal. In our second benchmarking scenario, we retained the original 3 features (2 predictive, 1 prognostic) and introduced additional varying numbers of features containing random noise (*n* = 7, 17, 37). Remarkably, the PBMF consistently outperformed VT with 7 (PBMF AUPRC = 0.834 ± 0.050; VT AUPRC = 0.746 ± 0.039) or 17 (PBMF AUPRC = 0.768 ± 0.044; VT AUPRC = 0.690 ± 0.040) random features (Fig. 2c). With 37 random features, both approaches exhibited similar performance (PBMF AUPRC = 0.650 ± 0.033; VT AUPRC = 0.644 ± 0.036).

We hypothesized that in noisy scenarios, the ensemble PBMF might incorporate suboptimal constituent models. Our third benchmark explored the impact of model pruning on enhancing ensemble performance. When employing only the top quartile (p75) or top decile (p90) models within the ensemble, we observed a marked improvement in PBMF performance, particularly in the presence of some (*n* = 7) or many (*n* = 37) random features (Fig. 2d). This pruning strategy outperformed VT, but it necessitated a larger ensemble (1024 versus 128) to achieve stable performance (Fig. 2d).

Our final benchmarking scenario investigated how the performance of the PBMF scales with the size of the training data set. In the simple case of 3 total features (2 predictive and 1 prognostic; i.e., benchmark 1), both the PBMF and VT methods exhibited diminished performance when training data were reduced from 1000 to 250 samples (Fig. 2e, Table S2). Despite this reduction, the PBMF still outperformed the VT (PBMF AUPRC = 0.786 ± 0.066; VT AUPRC = 0.752 ± 0.091). In the more complex scenario of 2 predictive, 1 prognostic, and 7 random features (i.e., benchmark 2), the performance of the PBMF matched or exceeded that of VT at all training data sizes tested (*n* = 250, 500, 1000, 2000, 4000; Fig. 2e). Although VT performance reached a plateau at 1000–2000 samples, the PBMF demonstrated continuous improvement and superior performance; notably, at the largest training data size tested (*n* = 4000), the PBMF (AUPRC = 0.967 ± 0.008) significantly outperformed the VT method (AUPRC = 0.788 ± 0.027). Lastly, the introduction of model pruning further enhanced PBMF performance at training data sizes greater than 500.

### PBMF identification of predictive biomarkers in across a diversity of clinical studies

Having established the success of the PBMF in simulated scenarios, we benchmarked the PBMF, VT, and SIDES across a diversity of 9 clinical studies, including real-world data, various cancer and non-cancer indications, and phase 1, 2, and 3 clinical trials. Overall, the PBMF markedly outperformed all other methods by consistently identifying predictive biomarkers (Fig. 3a). We detail the results of our benchmarking in the sections to follow.

**Fig. 3:**
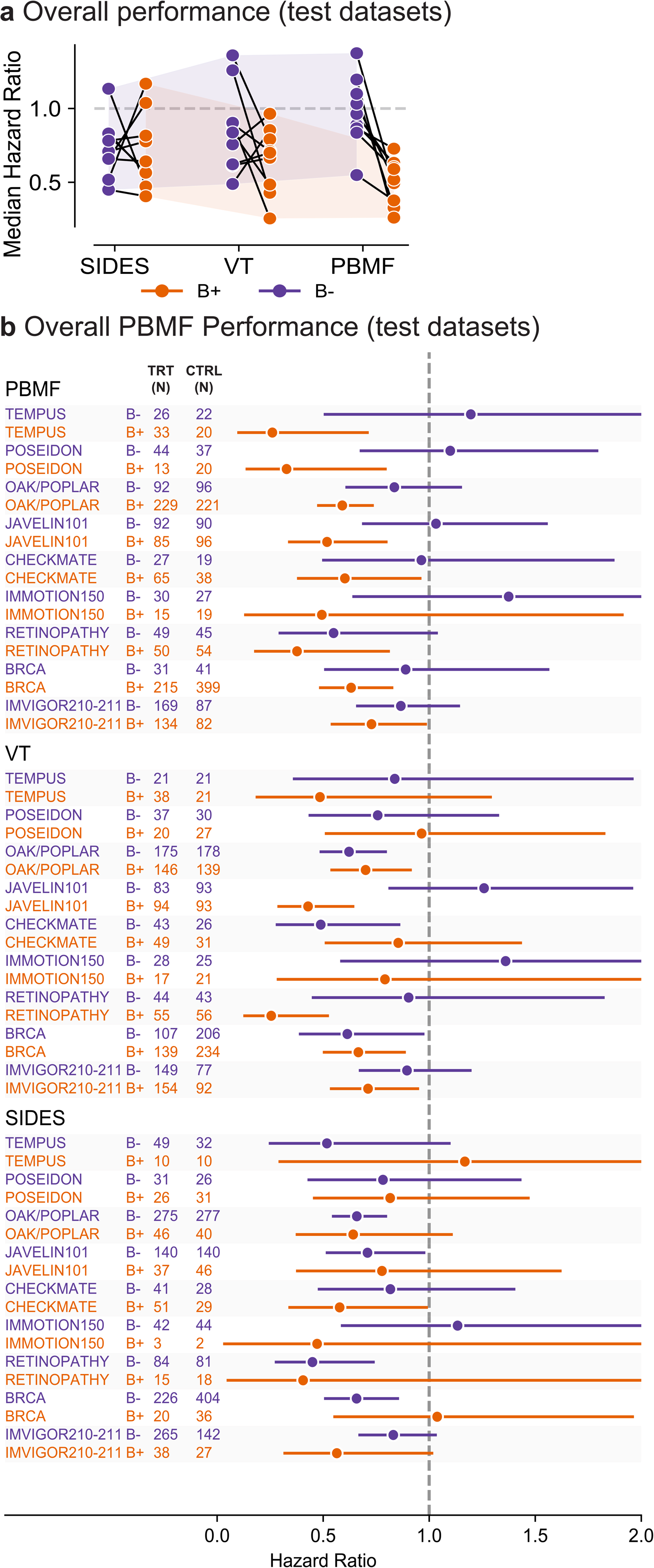
Evaluation of PBMF for predictive biomarker identification on real data sets against other methods. **a**, Hazard ratios for SIDES, VT, and PBMF methods across all 9 test datasets and across treatments for each biomarker status, B+ and B−. Points are connected if they represent hazard ratios computed for biomarker groups within the same dataset. Shaded areas correspond to the bounding box defined by the maximum and minimum hazard ratios for each method, for a given biomarker status, B+ and B−. **b**, Forest plot illustrating the performance comparison of PBMF with VT and SIDES methodologies, applied to test data sets. Shown are the hazard ratios and 95% confidence intervals from a Cox proportional hazards model fit to each treatment comparison within a biomarker status. Patient numbers (N) are shown to the left of the forest plot, where Trt = the treatment for which the predictive biomarker was desired (e.g. IO for TEMPUS) and Ctrl = the comparator treatment (e.g. chemotherapy for TEMPUS).

### Identification of predictive biomarkers in commonly used clinical datasets for survival analysis

We evaluated PBMF against VT and SIDES with well-characterized clinical datasets used in common practice for time-to-event statistical modeling (specifically survival analysis).^17,18^ We utilized breast cancer^19,20^ and diabetic retinopathy^21^ datasets, as these were the most feature-rich and appropriate for a predictive biomarker discovery task.

First, we benchmarked the PBMF against VT and SIDES for identifying a biomarker predictive of longer survival with hormone therapy + tamoxifen versus chemotherapy across the two available independent breast cancer data sets. Models were trained on the Rotterdam breast cancer cohort^22^ and subsequently tested on the German breast cancer study cohort.^19^ On the training data set, the PBMF (B+: hazard ratio [HR] = 0.71, confidence interval [CI] = 0.54–0.94, *P* = 1.69e-2; B−: HR = 1.91 CI = 1.48–2.48, *P* = 9.37e-7) and VT (B+: HR = 0.56, CI = 0.44– 0.70, *P* = 4.98e-7; B−: HR = 1.81, CI = 1.30–2.52, *P* = 4.32e-4) methods successfully identified a predictive biomarker, whereas SIDES found a prognostic biomarker (Fig. 3b, Fig. S1, Fig. 4a). On the test data set, only the PBMF generalized as a predictive biomarker (B+: HR = 0.63, CI = 0.48–0.83, *P* = 1.02e-3; B−: HR = 0.89, CI = 0.50–1.57, *P* = 6.84e-1), whereas both VT and SIDES were prognostic.

**Fig. 4:**
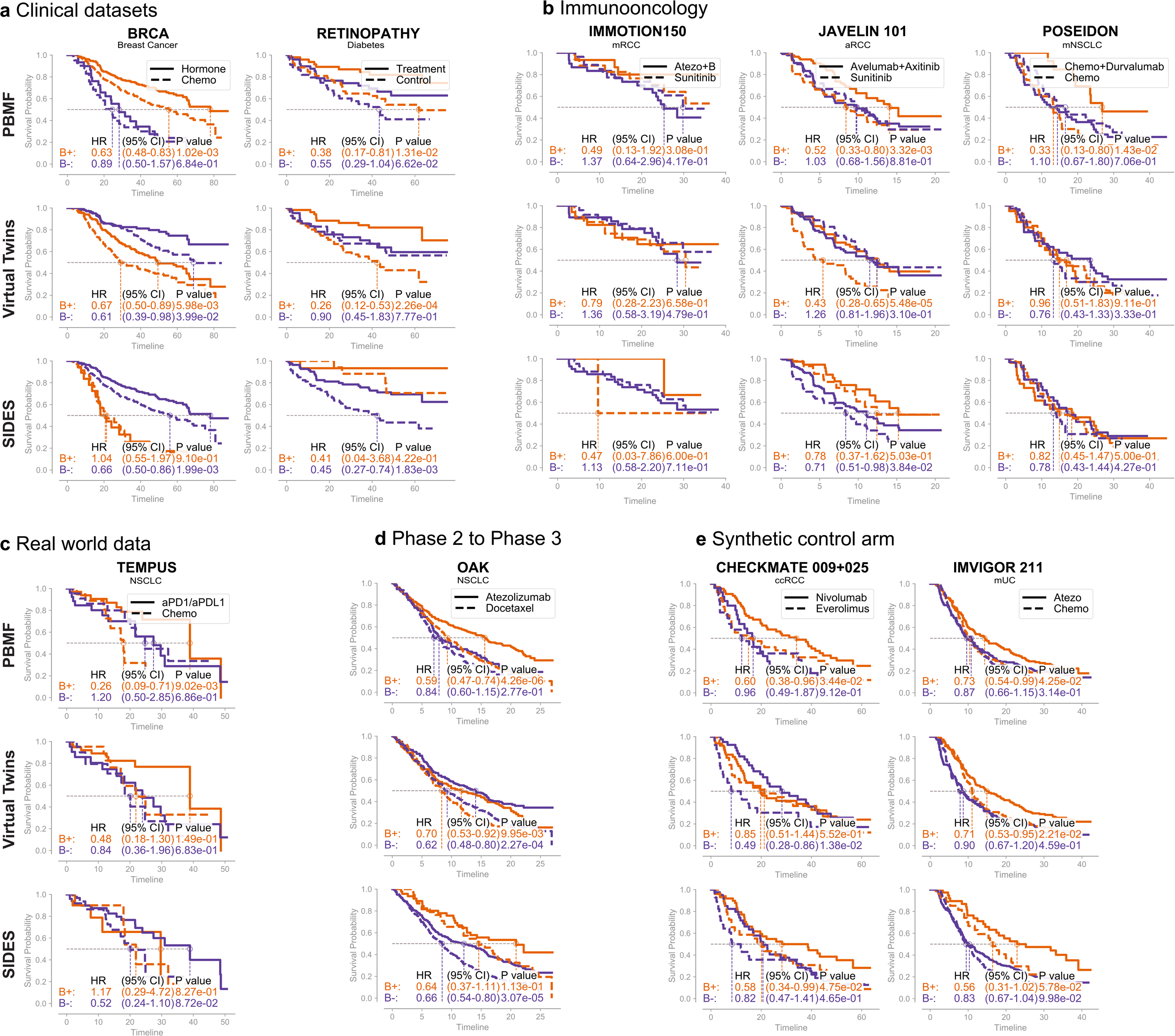
Kaplan-Meier curves for evaluation of PBMF for predictive biomarker identification on real data sets against other methods. Kaplan-Meier curves per treatment and biomarker status (from PBMF, VT, or SIDES), as evaluated on the **a**, test data from well-established clinical datasets for survival analysis (breast cancer and retinopathy), **b**, immunooncology clinical trial test data (IMmotion 150, JAVELIN 101, and POSEIDON), **c**, TEMPUS real-world data test set, **d**, OAK phase 3 clinical trial test data set, and **e**, clinical trial test data that utilized synthetic control arms (CheckMate 009 + CheckMate 0025 and IMvigor 211). Timeline is in months. Hormone, hormone therapy; chemo, chemotherapy; atezo, atezolizumab.

We next benchmarked the PBMF against VT and SIDES for identifying a biomarker predictive of longer time to vision loss with laser therapy versus no treatment in a study for treating diabetic retinopathy. On the training split of the data, the PBMF (B+: HR = 0.27, CI = 0.13–0.55, *P* = 3.67e-4; B−: HR = 0.69, CI = 0.38–1.24, *P* = 2.13e-1) identified the strongest predictive biomarker (Fig. S1). VT (B+: HR = 0.38, CI = 0.21–0.70, *P* = 1.88e-3; B−: HR = 0.55, CI = 0.28–1.09, *P* = 8.81e-2), and SIDES (B+: HR = 0.38, CI = 0.09–1.52, *P* = 1.71e-1; B−: HR = 0.46, CI = 0.29–0.74, *P* = 1.51e-3) found mostly prognostic biomarkers (Fig. S1a). In particular, for VT, the biomarker from the training data appears to enrich for reduced time to vision loss within each treatment, which is opposite to the desired behavior (Fig. S1b). This therefore discounts the otherwise favorable generalization of VT on the test split of the data (Fig. 3b, Fig. 4a). In contrast, the PBMF (B+: HR = 0.38, CI = 0.17–0.81, *P* = 2.26e-4; B−: HR = 0.55, CI = 0.29–1.04, *P* = 6.62e-2) identified a predictive biomarker, albeit with a prognostic component (Fig. 3b, Fig. 4a).

### Predictive biomarker identification in immunooncology

Encouraged by our promising results from simulated biomarker scenarios and well-established clinical datasets for survival analysis, we asked whether the PBMF would excel over VT and SIDES in the challenging predictive biomarker discovery space of immunooncology, specifically for immune checkpoint inhibitor (ICI) therapy. We trained and tested models on each of three phase 3 clinical trials (JAVELIN 101, NCT02684006; IMmotion 150, NCT01984242; POSEIDON, NCT03164616) for three different ICI therapies given in a first-line setting (avelumab, atezolizumab, durvalumab, respectively) for either renal cell carcinoma or non-small cell lung cancer (NSCLC). SIDES failed to find a predictive biomarker on the training data for IMmotion 150 and JAVELIN 101, whereas both the PBMF and VT consistently found a predictive biomarker on the training data for all three clinical trials (Fig. S1a).

On the test data for IMmotion 150, the PBMF trended the best towards a predictive biomarker, as it enriched for both for patients that had better survival across treatments within the B+ group (HR = 0.49, CI = 0.13–1.92, *P* = 3.08e-1), as well as across biomarker status within the ICI treatment (Fig. 4b). In contrast, although VT similarly trended towards a predictive biomarker (Fig. 3b), the B+ group across treatments trended towards worse survival than the B− group (Fig. 4b). When testing on JAVELIN 101, only the PBMF (B+: HR = 0.52, CI = 0.33–0.80, *P* = 3.32e-3; B−: HR = 1.03, CI = 0.68–1.56, *P* = 8.81e-1) generalized as a predictive biomarker. The PBMF identified a B+ group characterized by longer survivors in the avelumab + axitinib arm of interest versus all other groups and arms (Fig. 3b, Fig. 4b). Although VT appears to have found a generalizable predictive biomarker as well (B+: HR = 0.43, CI = 0.28–0.65, *P* = 5.48e-5; B−: HR = 1.26, CI = 0.81–1.96, *P* = 3.10e-1), examination of the Kaplan-Meier plots suggests that it instead identified a B+ group treated with the control therapy, sunitinib, that had worse survival versus all other groups and arms (Fig. 4b). Finally, when testing on POSEIDON, once again only the PBMF identified a predictive biomarker that can generalize (Fig. 3b, Fig. 4b; B+: HR = 0.33, CI = 0.13–0.80, *P* = 1..4e-2; B−: HR = 1.10, CI = 0.67–1.80, *P* = 7.06e-1).

In summary, PBMF demonstrated superior performance in all three phase 3 clinical trials for immune checkpoint inhibitor therapies, consistently identifying predictive biomarkers where SIDES failed and VT misidentified beneficial groups. PBMF reliably pinpointed patient groups with improved survival outcomes, highlighting its potential as a robust tool for predictive biomarker discovery

### Predictive biomarker identification with real-world data

Randomized controlled phase 3 clinical trials are often considered the gold standard for tasks like predictive biomarker discovery analysis. these datasets often take a significant amount of time to accumulate and require substantial investments. With the increasing availability of real-world evidence (RWE), we have chosen to benchmark PBMF against VT and SIDES despite challenges associated with the use of RWD, including issues related to inconsistent data quality, comparability, and bias.^23,24^ To facilitate this comparison, we curated a Tempus NSCLC real-world data cohort specifically to evaluate first-line ICI therapy versus chemotherapy (see methods for more details).

On the training data set, only the PBMF and VT yielded a biomarker with predictive value for ICI over chemotherapy, whereas SIDES exhibited a trend toward prognostic behavior (Fig. S1). On the test data set, only the PBMF (B+: HR = 0.26, CI = 0.09–0.71, *P* = 9.02e-3; B−: HR = 1.20, CI = 0.50–2.85, *P* = 6.86e-1) demonstrated enrichment for longer survivors specific to ICI therapy, indicating the discovery of a predictive biomarker that can generalize (Fig. 3b, Fig. 4c). In contrast, VT failed to generalize in the test data set (B+: HR = 0.48, CI = 0.18–1.30, *P* = 1.49e-1; B−: HR = 0.84, CI = 0.36–1.96, *P* = 6.83e-1), despite very strong predictive behavior observed in the training data set. The trend towards prognostic behavior failed to generalize for SIDES (B+: HR = 1.17, CI = 0.29–4.72, *P* = 8.27e-1; B−: HR = 0.52, CI = 0.24–1.10, *P* = 8.72e-2).

### Identification of individuals with improved survival outcomes to inform phase 3 trial design with early-stage clinical trial data

One critical application of predictive biomarker discovery is to inform the patient selection strategy for phase 3 clinical trials by using data from earlier phases. Building on the promising results from immunooncology and real-world data, we evaluated the PBMF against VT and SIDES in the context of representative clinical trial decision-making. Models were trained on clinicogenomic phase 2 trial data (POPLAR,^25^ NCT01903993), and tested on phase 3 trial data (OAK,^26^ NCT02008227). This evaluation aimed to determine which model could effectively guide patient selection for second-line atezolizumab therapy versus chemotherapy in NSCLC (i.e., the OAK trial), relying solely on data from earlier studies.

Both PBMF (B+: HR = 0.30, CI = 0.19–0.48, *P* = 2.57e-7; B−: HR = 2.41, CI = 1.41–4.11, *P* = 1.25e-3) and VT (B+: HR = 0.38, CI = 0.24–0.60, *P* = 3.72e-5; B−: HR = 1.14, CI = 0.72–1.78, *P* = 5.76e-1) identified a predictive signal from the phase 2 POPLAR training data. SIDES identified a mixed predictive and prognostic signal (B+: HR = 0.42, CI = 0.14–1.21, *P* = 1.08e-1; B−: HR = 0.75, CI = 0.54–1.05, *P* = 9.51e-2) (Fig. S1). Importantly, when the three models trained on POPLAR study data were applied as a hypothetical patient selection biomarker for the phase 3 OAK trial test data, only the PBMF generalized as a predictive biomarker (Fig. 3b, Fig. 4d; B+: HR = 0.59, CI = 0.47–0.74, *P* = 4.26e-6; B−: HR = 0.84, CI = 0.60–1.15, *P* = 2.27e-1).

Both VT (B+: HR = 0.70, CI = 0.53–0.92, *P* = 9.95e-3; B−: HR = 0.62, CI = 0.48–0.80, *P* = 2.27e-4) and SIDES (B+: HR = 0.64, CI = 0.37–1.11, *P* = 1.13e-1; B−: HR = 0.66, CI = 0.54– 0.80, *P* = 3.07e-5) yielded only prognostic biomarkers (Fig. 3b, Fig. 4d). Compared with the biomarker-evaluable population (BEP) in the OAK trial (Fig. S4), the PBMF B+ subpopulation yielded a ∼9% decrease in risk of death for atezolizumab versus docetaxel treatment (PBMF, HR = 0.59; OAK BEP HR = 0.65). Thus, to hypothetically inform strategies for patient selection in phase 3 clinical trials, only the PBMF successfully identified a predictive, high-prevalence biomarker from phase 2 data that generalized to phase 3 results.

### A discovery pipeline for predictive biomarker prototypes

Given the consistent ability of the PBMF to identify a predictive biomarker, particularly in clinical trial settings, we devised an end-to-end biomarker discovery pipeline that generates a human-understandable predictive biomarker prototype, poised for translation into clinical settings (Fig. 5a). We utilized the PBMF ensemble-pruned model described in the preceding section (Fig. 3b, Fig. 4d), which was trained solely on phase 2 clinical trial data (Fig. 5b), to identify a predictive biomarker (Fig. S2a–c, Methods). Utilizing a consensus score across the models within the ensemble, we determined an optimal biomarker probability score cutoff to classify B+ and B− samples, subsequently referred to as pseudo-labels (Fig. 5d, Methods). These pseudo-labels were then used for the distillation of the complex neural network original PBMF model into a simple interpretable model—a decision tree—that could inform a strategy for a clinical study (Fig. 5d, Fig. S2a–c, Methods).

**Fig. 5:**
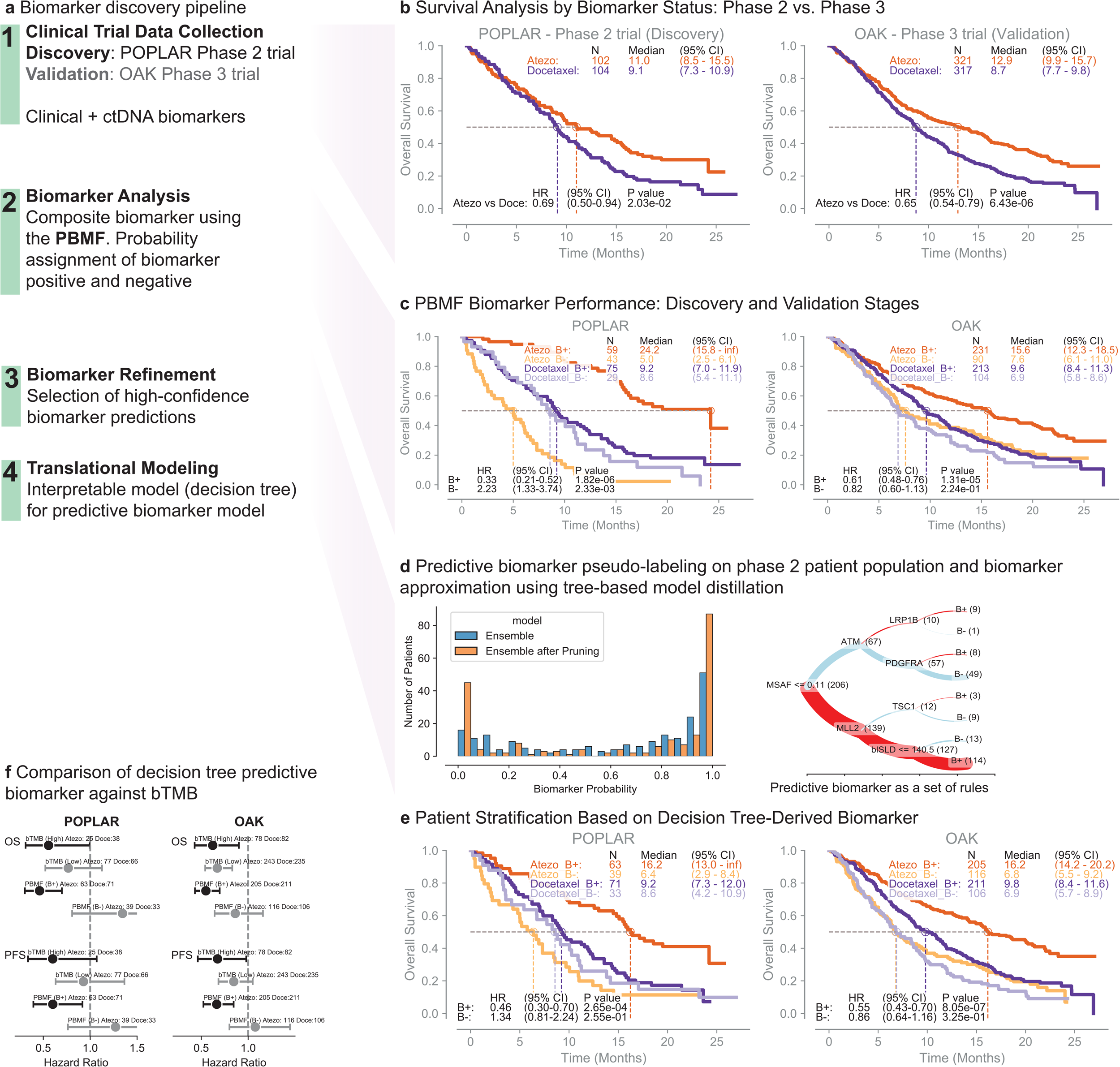
Application of PBMF in the design of biomarker-driven clinical trials. **a**, Overview of the proposed integrative framework for the discovery of predictive biomarkers in phase 2 trials to enhance phase 3 trial design, incorporating initial data acquisition from early-phase trials, PBMF analysis, biomarker optimization through interpretable models, and subsequent application in clinical trial planning. **b**, Clinical trial data and endpoints collection: Kaplan-Meier curves for the discovery (POPLAR phase 2 clinical trial) and the test (OAK Phase 3) data sets. **c**, Identification of predictive biomarker: using the discovery data set (POPLAR trial) the PBMF successfully finds a biomarker that identifies which patients will survive longer on atezolizumab but not docetaxel. This biomarker generalizes to the OAK trial test data. **d**, Refinement of predictive biomarker: the enhancement of the predictive biomarker involves pruning to eliminate spurious models from the ensemble (left) and the subsequent derivation of a rule set that encapsulates the biomarker’s predictive power (right). Red lines, B+; blue lines, B−; line thickness is proportional to number of patients in parenthesis. **e**, Patient stratification using the simplified predictive biomarker identified in the POPLAR trial and subsequently applied to the OAK trial. **f**, Comparison of the predictive biomarker against blood TMB in the discovery (POPLAR) and test (OAK) data sets, with an additional evaluation of the biomarker on progression-free survival (PFS), despite the PBMF’s initial training on overall survival (OS). Numbers of patients is shown for each treatment and biomarker status. Shown are the hazard ratios and 95% confidence intervals from a Cox proportional hazards model fit to each treatment comparison within a biomarker status. Atezo, atezolizumab; Doce, docetaxel.

### Use of knowledge distillation from the PBMF neural network to produce a simple decision tree with improved predictive value

Similar to the original PBMF from which it was derived, the distilled decision tree PBMF biomarker was predictive on both the phase 2 trial training data (B+: HR = 0.46, CI = 0.3–0.7, *P* = 2.6e-4; B−: HR = 1.34, CI = 0.8–2.2, *P* = 0.2) and phase 3 trial test (B+: HR = 0.55, CI = 0.43– 0.7, *P* = 8.05e-7; B−: HR = 0.86, CI = 0.64–1.16, *P* = 0.3) data sets (Fig. 5e). Importantly, the HR of the distilled decision tree was improved by approximately 7% compared with the original PBMF (original PBMF HR = 0.59; distilled decision tree PBMF HR = 0.55; see Fig. 5c, e), owing to the reduction in prevalence from 80% to 64%. Notably, the original PBMF had a ∼9% decrease in risk of death within the B+ atezolizumab versus docetaxel-treated subpopulation relative to the BEP in the OAK trial, and the distilled decision tree PBMF had a ∼15% decrease in risk of death (distilled PBMF HR = 0.55; original PBMF HR = 0.59; OAK BEP trial-reported HR = 0.65, OAK intent-to-treat HR = 0.73).

Upon scrutinizing the decision tree of the distilled PBMF, we observed that the predictive biomarker comprises a specific subset of clinical and genomic features: the maximum circulating tumor DNA ctDNA allele frequency (MSAF), sum of longest diameter of target lesions at baseline (blSLD), and mutation status on the *MLL2, TSC1, ATM, PDGFRA* and *LRP1B* genes (Fig. 5d). Collectively, all these features drive the predictive nature of the biomarker. With the exception of *ATM* mutations, which were both predictive and prognostic (POPLAR: mutation [Mut] B+ HR = 0.33, wild type [Wt] B− HR = 0.776; OAK: Mut B+ HR = 0.43, Wt B− HR = 0.68) but with a notably low prevalence (28 patients for *ATM* B+/Mut and 205 for the distilled PBMF B+), each individual feature fell short in matching the biomarker prevalence or the consistent, predictive signal of the collective (Fig. S3, Table S3). Furthermore, in comparison with a commonly described single-feature ICI biomarker, blood TMB,^27-29^ the PBMF more robustly enriched for longer survival for both the training and test clinical trial data sets (Fig. 5e, f; Table S4).

### Predictive biomarker discovery with synthetic control arms

Early phase trials are often single-arm studies, complicating efforts to derive biomarkers specific to a treatment of interest. Recent FDA guidance suggests common^30^ or external^31^ control arms might be used in certain settings to minimize redundancy, especially for and motivated in large part by Oncology drug discovery. We therefore evaluated our approach in this ‘synthetic control arm’ scenario, whereby we used a fraction of phase 3 control arm data exclusively for model training alongside phase 2 single-arm trial data.

In the context of pre-treated advanced clear cell renal carcinoma (ccRCC), PBMF, VT, and SIDES all identified a predictive biomarker for ICI therapy on the training data from the nivolumab arm of phase 2 CheckMate 010 (NCT01354431) and a synthetic control arm from a random subset of patients receiving everolimus from phase 3 CheckMate 025 (NCT01668784; Fig. S1). However, only the PBMF generalized to the test dataset on the combined population from phase 1 CheckMate 009 (NCT01358721) and phase 3 CheckMate 025 trials (Fig. 3b, Fig. 4e; excluding those from CheckMate 025 used for training; B+: HR = 0.60, CI = 0.38–0.96, *P* = 3.44e-2; B−: HR = 0.96, CI = 0.49–1.87, *P* = 9.12e-1). SIDES trended towards a prognostic biomarker (B+: HR = 0.58, CI = 0.34–0.99, *P* = 4.75e-2; B−: HR = 0.82, CI = 0.47–1.41, *P* = 4.65e-1), whereas VT did not generalize, as it displayed a predictive biomarker for the control arm (B+ HR = 0.85, CI = 0.51–1.44, *P* = 5.52e-1; B−: HR = 0.49, CI = 0.28–0.96, *P* = 1.38e-2).

Overall, the PBMF identified a B+ subpopulation with a 12% decrease in risk of death when treated with nivolumab versus everolimus, relative to the BEP in the combined CheckMate 009 and 025 trials (Fig. 3b, Fig. S4; PBMF HR = 0.60; CheckMate 009 and 025 BEP HR = 0.68; CheckMate 025 BEP trial-reported HR = 0.69; CheckMate 025 intent-to-treat HR = 0.73).

The PBMF also generalized well in an additional independent cohort examining atezolizumab versus chemotherapy in locally advanced or metastatic urothelial carcinoma (mUC). In this analysis, we included all available input features at baseline (Age, sex, ECOG, pIL-8 expression, and liver metastasis) and on-treatment (pIL-8 after 6 weeks) to evaluate their association with overall survival. On the training data from the atezolizumab arm from phase 2 IMvigor210 (NCT02951767, NCT02108652) and a synthetic control arm from a random subset of patients receiving chemotherapy from phase 3 IMvigor211 (NCT02302807), only the PBMF and VT but not SIDES yielded a biomarker with predictive value of atezolizumab over chemotherapy (Fig. S1). Similarly, on the test dataset (IMvigor 211 excluding patients used for the training synthetic control arm), both PBMF (B+: HR = 0.73, CI = 0.54–0.99, *P* = 4.25e-2, B−: 0.87, CI = 0.66– 1.15, *P* = 3.14e-1) and VT (B+: HR = 0.71, CI = 0.53–0.95, *P* = 2.21e-2; B−: HR = 0.90, CI = 0.67–1.20, *P* = 4.59e-1) generalized well as a predictive biomarker (Fig. 3b, Fig. 4e). This corresponded to a 10% and 12% decrease in risk of death, respectively, when treated with atezolizumab versus chemotherapy, relative to the BEP in the IMvigor 211 trial (Fig. 3b, Fig. S4; PBMF HR = 0.73; VT HR = 0.71; IMvigor 211 BEP HR = 0.81; IMvigor 211 intent-to-treat HR = 0.85).

## DISCUSSION

Across diverse, challenging benchmarks spanning simulated scenarios through informing strategies for patient selection in clinical trials, the PBMF outperformed other methods for discovering predictive biomarker signals. Among comparator methods, only the PBMF found signals that were consistently predictive across training and test data sets. Along with the PBMF’s ability to accurately identify known IO biomarkers from phase 2/3 trials, we also showed that the PBMF can nominate a novel composite biomarker from a set of clinicogenomic features that outperformed blood TMB.

We emphasize here the importance of the predictive constraint embedded in the PBMF. A common pitfall in biomarker discovery is to focus only on identifying populations with enhanced responses to a specific treatment.^32^ In these cases, one cannot distinguish between a biomarker that is prognostic versus one that enriches for better responses specifically in a treatment of interest. Thus, the PBMF loss function enforces the constraint that a biomarker must be considered in the context of a control treatment.

Beyond its contrastive loss function, the PBMF stands out as a unique end-to-end API for predictive biomarker discovery. The results presented here underscore the superior performance of an ensemble PBMF consisting of fully connected neural networks. At the same time, our API is versatile and compatible with any differentiable model. This flexibility makes it possible to explore predictive biomarker signals using input features from single or multiple modalities, or diverse data representations, including various combinations thereof. For instance, an attention-based transformer model could effectively model unstructured data such as clinical notes. This opens the door to leveraging pretrained models, e.g. large-language models or other patients’ embeddings derived from foundation models, to imbue the PBMF with prior knowledge, potentially enabling successful predictive biomarker discovery even in situations with limited or noisy data.^33^ Lastly, the PBMF provides tools to refine a biomarker toward a particular downstream application, i.e., prevalence constraints, simulations, and knowledge distillation, for clinical deployment.

In our patient selection strategy example, we successfully distilled a complex ensemble neural network model into a simple decision tree. In this regard, we can view the PBMF as a highly effective search function, as we required the complex model to discern whether a predictive signal exists and what features may drive it. Alternatively, one could model patient risk through a multivariate Cox PH model with interaction terms for treatment. Although this approach may theoretically achieve similar results, it may be impractical to implement. Whereas the gradient descent within the PBMF will implicitly traverse the vast expanse of potential feature combinations and interactions, one would have to systematically and explicitly test every single potential case when using a Cox PH model. Further, the PBMF accounts for treatment effects simultaneously within its loss function, whereas a Cox PH model requires enumeration of each hypothesized treatment-feature interaction.

We concede that there are limitations of the PBMF, although most are common to any biomarker nomination process. First, there is no guarantee that a predictive signal exists amongst the available features in a given cohort. Indeed, many well-established clinical datasets for survival analysis contain only age and/or sex features, and only prognostic biomarkers can be found with any modeling approach. Related, with the known challenge of limited data sets and high heterogeneity in patient populations, the PBMF cannot be used to determine whether the data are adequate and representative of the target population and biology. Nevertheless, it is noteworthy that the PBMF demonstrated superior performance in scenarios with small data sizes. In situations with substantial data, PBMF scaled with data size, whereas the performance of the VT method reached a plateau. Second, the ensemble PBMF may be unable to maintain its magnitude of predictive power when distilled into a simple model, as there is often a tradeoff between a biomarker’s predictive power and its parsimony.^34^ However, the enhanced interpretability of the model may contribute to a better understanding of the biological factors underpinning the predictive signal of the biomarker. More generally, with any biomarker nomination process, there is the risk of overfitting to the training data and lack of generalization when the biomarker is deployed prospectively. Encouragingly, at least within the scope of the current study, the PBMF provided concordant results between training and test sets and to a greater degree than the comparator methods. Third, while the PBMF outperformed other methods in discerning predictive signals from noisy or prognostic features, we might still find that strongly prognostic features can impede the identification of predictive signals, and therefore our method could potentially gain more from prior feature selection. Fourth, the PBMF’s contrastive loss function formulation tends to attenuate the discovery of biomarkers that show a modest positive effect in the control treatment but a more substantial benefit in the treatment of interest. Finally, the PBMF is a discovery tool, and any biomarker hypothesis requires prospective clinical validation.^35-37^

Specific considerations and limitations apply when using any predictive biomarker method to inform late-stage clinical trial decision-making. As alluded to earlier, data availability is often limiting. The success of the PBMF in identifying potential predictive biomarkers from real-world data and from using synthetic control arms is thus promising. Future work will be required to know whether synthetic control arms from non-randomized evidence (i.e. real-world data) could be used; any such exploration would need to carefully consider the substantial heterogeneity within patient populations. A related point is that it is often difficult to ensure that cohorts are comparable across studies, as the intent-to-treat clinical trial design guarantees only within-trial comparisons. Moreover, considering the rising trend of combination therapies, it will be crucial to investigate the PBMF’s performance across various arms and their pairwise combinations. As our study is retrospective in nature, an important next step would be to validate the PBMF prospectively in a future clinical study. Finally, future work can explore the tradeoff between data maturity, ability to extract a predictive signal, and phase 3 trial investment decision timing. Our benchmarks nonetheless demonstrate that with the availability of the appropriate data, the PBMF could nominate a predictive biomarker that is likely to outperform the original study design in selecting patients who would derive greater benefit from the new treatment in a phase 3 study. The use of the PBMF has the potential to improve strategies for patient selection over what can be achieved with conventional study designs.

## METHODS

### Predictive biomarker loss function

The PBMF (Fig. 1) uses as input time-to-event data with censoring, a treatment label, and a feature matrix (*n* patients by *f* features). The feature matrix X ∈ □^*f*^ is used as the input to a fully connected neural network of user-defined depth and width.

The goal of the neural network is to assign patients to either the B+ or B− group. To refine this categorization, we employed a contrastive learning approach in which patients in the B+ group, when under treatment, show an improvement in survival times compared with those in the B− group. Conversely, in the control arm, the model aims to minimize the differences in survival times between the two biomarker groups according to the principle of contrastive learning.^38-40^

The distinction or similarity in survival times is quantified using log-rank test statistics^41^ within each treatment arm as follows:

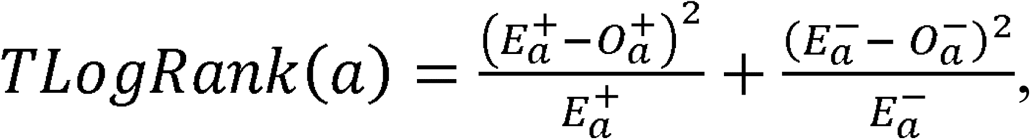

where the 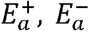 pair represents the expected number of events for the treatment *a*, under B+ and B−, respectively. The 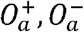 pair depicts the observed events within the treatment *a* for B+ and B−, respectively.

Formally, the expected and observed events are defined as follows:

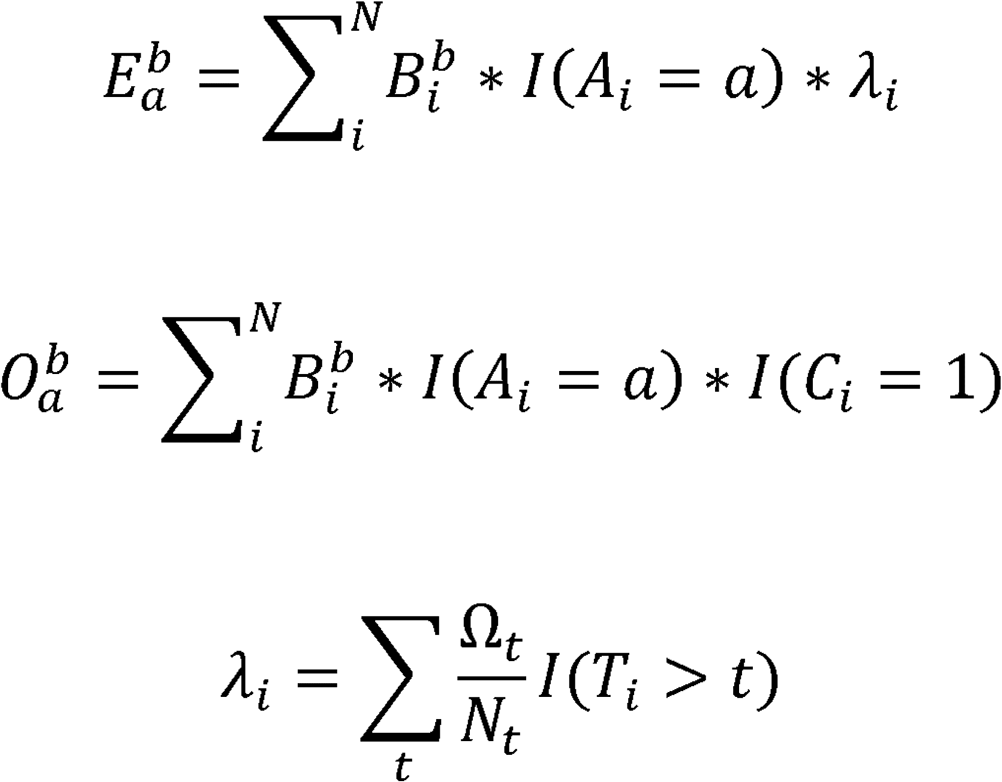

where the treatment arm is defined by *a* ∈{*Treatment* (*Tr*), *Control* (*CR*)} and the indicator function *I*(*A*_*i*_ = *a*) determines whether the patient *i* is under treatment *a* or not. The biomarker group is defined by the output of the neural network where *b* ∈ {positive (+), negative (–)}. Therefore, each patient *i* has a probability of being labeled as being in the positive 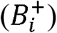 or negative 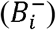 group. *C*_*i*_ represents the censoring status of patient *I*, and *λ*_*i*_ is a scalar independent on the parameters of the neural network and can be precalculated (see Meier et al.^42^). Ω_*t*_ is the number of observed events at time *t*, and *N*_*t*_ is the number of subjects at risk at time *t*.

The log-rank test for the treatment and control is then defined as:

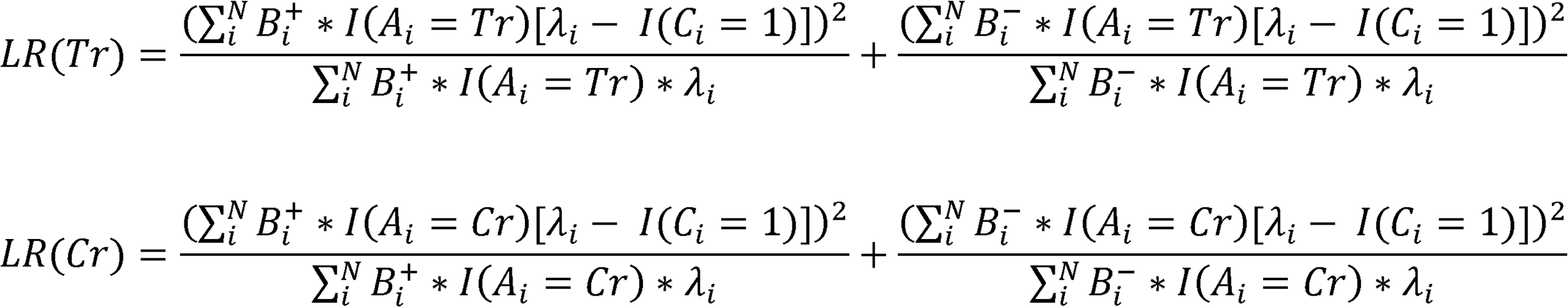

The contrastive nature of the loss function is evident in its formulation as follows:

- Treatment arm optimization: For patients receiving the actual treatment, the model maximizes the survival time difference between B+ and B− groups. This is quantified by the treatment log rank test score, *LR*(*Tr*).
- Control arm optimization: For the control group, the model minimizes the survival time difference between the two biomarker groups. This is quantified by the control log rank test score, *LR*(*Cr*).

The contrastive loss for the predictive biomarker is then defined as the ratio between the control log rank test score by the treatment log-rank test score:

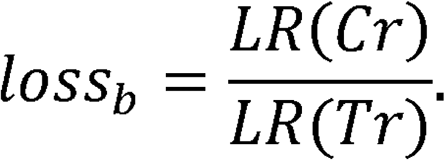

The custom contrastive loss is the ratio of two log-rank tests computed over the time-to-event data, grouped by the treatment label, and stratified by the neural network output score. During optimization, the neural network learns a set of parameters that outputs scores to maximize the separation (i.e., larger log-rank test statistic) for the treatment while minimizing the separation (i.e., smaller log-rank test statistic) for the control. This ensures that the neural network will learn to generate a predictive biomarker score, since it will only stratify patients for a specific treatment.

We also integrated a population prevalence term to the loss to enable the model to identify a predictive biomarker given a specific desired minimal population (*minP*) such that:

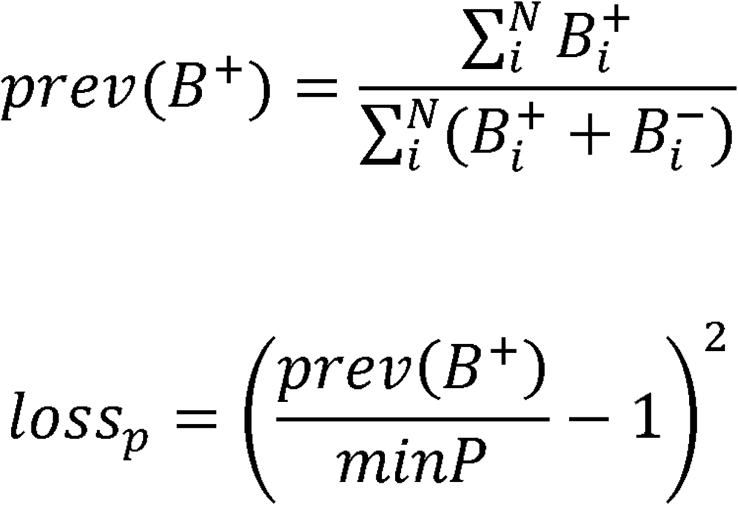

The *loss_p* will have a minimum value of 0 when *minP* is equal to the population of *B*^*+*^. Finally, the composite PBMF loss function takes the following form:

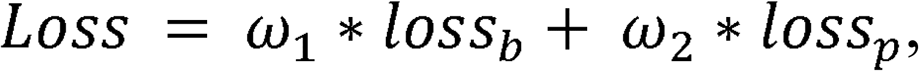

where *ω*_1_ and *ω*_2_ dictate the contribution of each loss component. For example, when *ω*_2_ = 0, the PBMF finds a population with the best predictive power independent of the number of patients, and when *ω*_2_ = 0.5 the PBMF identifies a predictive biomarker of the treatment at a 50% patient prevalence.

### Biomarker scoring

The output of the neural network (B ∈ □^2^) is composed of two units representing the B+ and B− scores {*b*^+^, *b*^−^}. Scores are then passed through a SoftMax activation to convert the network scores into probabilities. Thus, the biomarker scores for a given patient *i* can be expressed as:

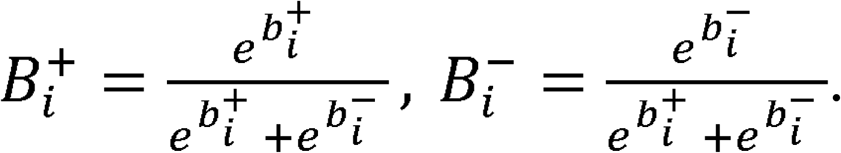

The probability of the negative biomarker can be written as *B*^−^ = (1 – *B*^+^). In this way, *B*^+^ values close to 0 indicate B− and values close to 1 indicate B+. We assume the B+ to be contained within the neuron at index 0 from the output of the neural network. However, because the loss function does not have control of the directionality of the assignments, B+ can be arbitrary placed in neuron at the index 0 or 1. Therefore, after training and when making predictions, we corrected the B+ by computing the HR between the B+ and B− within the treatment arm as 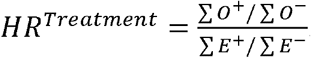. Thus, an *HR*^*Treatment*^ < 1 defines the B+ in the neuron 0, whereas an *HR*^*Treatment*^ > 1 defines the biomarker positive in the neuron 1.

With ensemble of neural networks, for a given patient and a total of *M* neural network models, we generated a set of scores 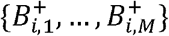 and computed a consensus score defined by the average score over all the models in the patient *i* such that 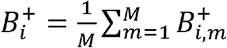.

### Feature and patient subsetting during model training

A random subset of patients and features can be specified (Table S1) to guard against model overfitting. Patient subsetting is performed before model loss computation, and a different subset of patients will be excluded at each gradient update. Feature subsetting is performed before model training, and the given model will only train on the feature subset; when training an ensemble, each model will utilize its own unique random subset. During ensemble model evaluation, no patients or features are excluded.

### PBMF ensemble model pruning

Under the assumption that some models in the ensemble perform poorly and damage the entire ensemble’s performance, we implemented the following model pruning approach. We first binarized the set of scores, 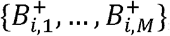 generated from the trained ensemble, using the default 0.5 score threshold for the PBMF. Using this *N* patients by *M* models binary matrix, *R*, we then compute an *N* × *N* patient agreement matrix, *A*, by calculating the proportion of models that assigned two different patients to the same class^43^:

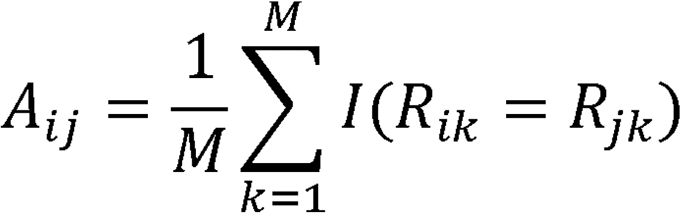

*A* contains 1 along its diagonal, is symmetric, and contains values ∈ [0,1]. Patients with similar scores across each model in the ensemble will tend to have higher values; those with dissimilar scores will have lower values. Each column or row of *A* represents how consistently patients were assigned to a particular class by the models in the ensemble, from the reference point of one patient.

We then computed the Pearson correlation between each column in *A* with each column in *R* to generate an *N* × *M* matrix, *C*, of correlation coefficients that represents how well the patient scores from an individual model in the ensemble correlate with the patient agreement matrix. We assumed that only a minority of models have poor performance, such that we should keep models that agree on how patients should be scored and discard models that disagree. This was done by selecting a percentile, e.g., the 90th percentile of all the correlations. By thresholding on the value in *C* associated with this percentile, the models were sorted by the number of times that each model exceeded the threshold, to generate a 1 × *M* vector of counts. We then thresholded on the value associated with our percentile in this vector to return the final subset of models, *M*_*S*_, that exceed this threshold. A new consensus score was then computed as the average score across the reduced set of models in the ensemble.

### Model distillation: pseudo-labeling

The distribution of scores generated from the ensemble is used to identify patients with “high-quality” predictions, i.e., those whose distributions are heavily skewed toward 0 (strongly B−) or 1 (strongly B+).

To identify the patients with the best high-quality scores, we choose a 0.5 cut point and add an offset value ε, such that the biomarker label for a patient *ii* is defined as:

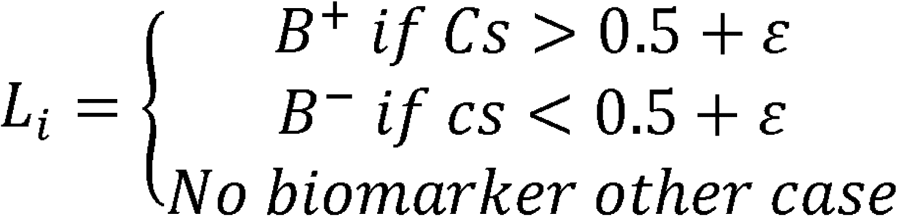

We set ε ∈ {0, 0.1, 0.2, 0.3, 0.4} and then fitted a Cox PH model to compute the hazard ratios between the treatment and the control arms for both the B+ and B−. The optimal ε score is extracted by determining the maximum difference between the absolute log of the B+ and B− hazard ratios.

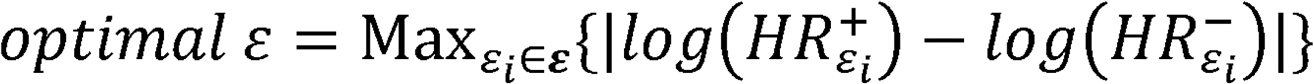

We then applied the optimal ε to compute a reduced set of patients with high-quality scores.

### Model distillation: tree-based model explainability

Once the high-quality population is defined, a tree classifier (python sklearn^44^ tree classifier package, max_depth = 3, random_seed = 0) is fit, using the input features and the B+ and B− as the labels. The goal of the tree classifier is to define a simple rule that approximates the neural network–derived predictive biomarker. The tree model was then applied to the test data sets.

### VT implementation

We implemented the VT approach proposed by Foster et al.^15^ as follows. We used a random survival forest model^45^ to predict time-to-event based on the log-rank test loss (pySurvival^46^). We built two survival models {*M*_*T*_, *M*_*C*_}, where *T* and *C* refer to the population under treatment and under the control, respectively. Each model was trained using only its respective population. We then computed the difference in risk score between the treatment and control models to define the counterfactual risk score *r*_*i*_ = *M*_*T*_(*i*) – *M*_*C*_(*i*) for any given patient *i*.

To stratify patients into B+ and B−, we computed the median value of the counterfactual risk score distribution across all patients and assigned to B+ those patients below the median score (low risk) and to B− those with a counterfactual risk score above the median. Consequently, this design choice intrinsically classified patients evenly, 50% being assigned to B+ and the remaining 50% to B−. This can potentially lead to an overestimation of favorable results in data sets where the predictive biomarker prevalence is 50%.

For simulations, model hyperparameters were tuned as described in Supplemental Information and Table S5. Model hyperparameters for identifying predictive biomarkers for clinical studies is described in Table S1.

### SIDES implementation

The SIDES algorithm was set for survival analysis using the time and event features as the targets and the treatment versus control setting. The features used were the same as those used for PBMF and VT and depended on the analyzed data set. We used the R implementation of SIDES provided by the SIDES authors (sides.dylib, CSIDES.r, and stochSIDES_util.R). We selected the best biomarker sorted by the adjusted *P* value and assigned it as B+. The discovered predictive biomarker rule was then validated in a given independent test set. Model hyperparameters for identifying predictive biomarkers for clinical studies is described in Table S1.

### Synthetic data generation

We generated 10,000 patients for each data set. For a given replicate, 2000 patients (20%) were randomly selected, without replacement. Among those selected, a 50-50 training/test split was performed. Evaluation metrics are reported only from the test set. Proportional hazard assumptions were imposed to induce each one of the behaviors (Fig. 2a). The ability of each methodology to correctly call the biomarker was measured by recording the precision, recall, and AUPRC of a holdout test data set (2000 patients for each data set).

The generation of synthetic data sets involves three stages. Initially, a set of covariates with predetermined level of correlation and prevalence is defined (Fig. 2a). These covariates establish subgroups for which desired hazard ratios will be generated. For the parametric model, the cumulative hazard is

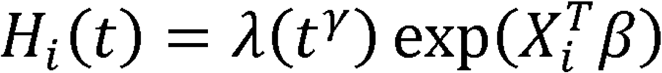

Where *X*_*i*_ is a vector of covariates associated to the parameters β. The β parameters used to sample survival times can be estimated after setting the HR requirements between groups. For example, assuming a treatment variable and a predictive biomarker, we can define the following hazard ratios:

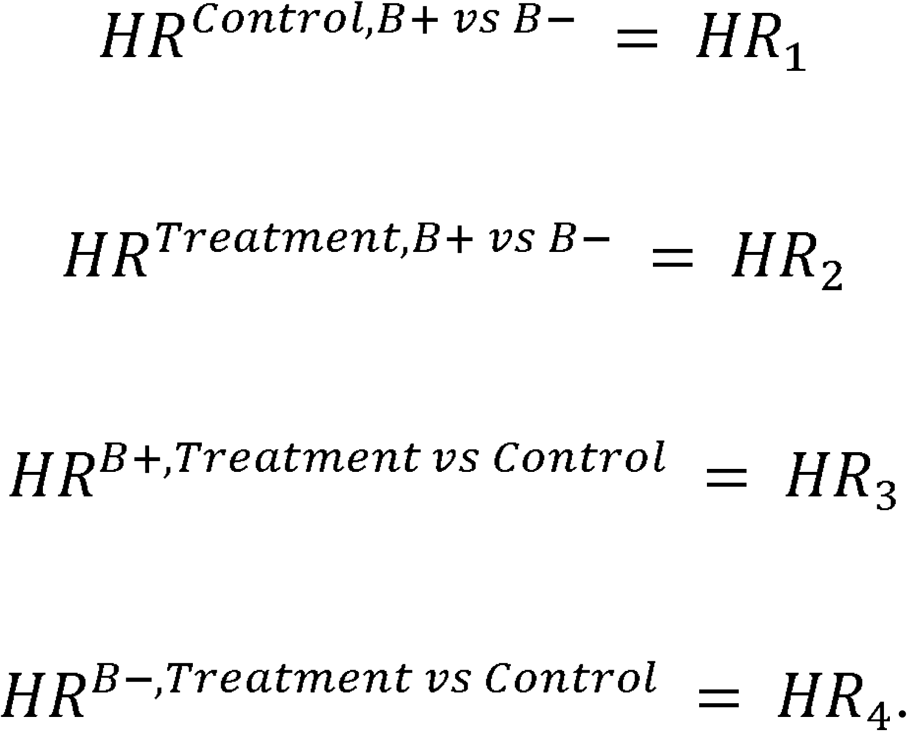

The time-independent part of *H*_*i*_ (*t*) can be expanded as:

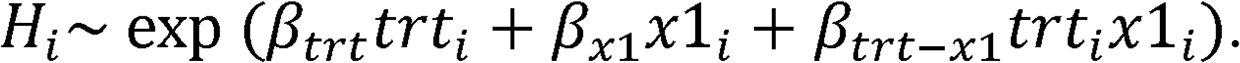

Replacing for each one of the cases in equation 1, we obtain the following equations:

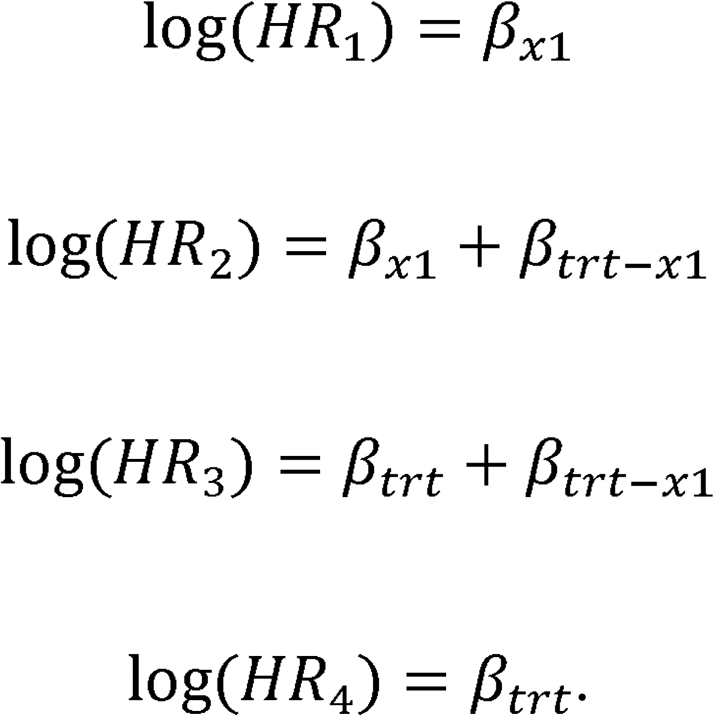

Random survival times are then obtained using the technique outlined in Crowther and Lambert (2013),^47^

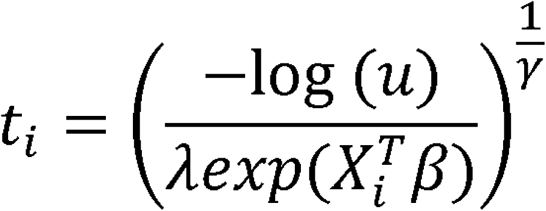

where *λ* and γ and are the scale and shape parameters, and *u* is a random variable sampled from the uniform distribution *U*(*0, 1*). Note that additional censoring, not covered in this work, can also be introduced.

### Real-word and clinical data sets

Hyperparameters (Table S1) were tuned for the PBMF, VT, and SIDES for each clinical dataset, using only training data.

The Rotterdam breast cancer cohort^20^ (863 patients) was used as a training data set, and the German breast cancer study cohort^19^ (686 patients) was used as a test data set. We selected only patients treated with hormone-based treatments and chemotherapy. The 7 features used for training the PBMF are age, menopause, tumor size, tumor grade, number of nodes, pr (progesterone receptor status), and er (estrogen receptor status). We trained the model using overall survival and death.

The DIABETIC retinopathy study^21^ evaluates the treatment of laser coagulation to delay diabetic retinopathy. In this study, 197 patients underwent treatment in one eye, while the other eye remained untreated. The treatment eye, right or left, was randomized. Treating each eye as an individual sample resulted in 394 observations in the dataset. The event of interest was the time from the start of treatment to the time when visual acuity dropped below 5/200 for two visits in a row. Censoring was caused by death, dropout, or the end of the study. Age, diabetes type, and risk score were included as the features of this dataset. Diabetes type was a binary feature indicating juvenile diabetes (diagnosis before age 20) or adult. Risk score was defined by the Diabetic Retinopathy Study, and a score greater than 6 out of 12 indicates high risk. The dataset was split into training and testing at a prevalence of 50% (random seed = 0).

The randomized phase 2 clinical trial IMmotion150 evaluated the efficacy of atezolizumab (anti-PD-L1) alone or in combination with bevacizumab (anti-VEGF) versus sunitinib (RTK inhibitor) in treatment-naive metastatic renal cell carcinoma (mRCC). Data from IMmotion150 was downloaded from Yuen et al.^48^ and comprised a total of 248 patients with no missing values (84 atezolizumab, 81 sunitinib, and 83 atezolizumab + bevacizumab). Available features on this dataset: age, sex, liver metastasis, previous nephrectomy, T-cell effector signature score, Plasma IL8, SLD (sum of longest tumor diameter) and sample type (primary / metastatic). IMmotion150 dataset was split into training / testing with a 50% prevalence, stratified by treatment and overall survival event (random seed = 0). The PBMF was trained to discriminate between atezolizumab + bevacizumab against sunitinib using overall survival time and event as endpoints (Fig. 3).

The JAVELIN Renal 101 trial evaluated the effectiveness of avelumab (PD-L1) plus axitinib (chemotherapy) versus sunitinib in advanced renal cell carcinoma (aRCC). Clinical response, PD-L1 status and RNA derived signatures (pathway scores) were downloaded from the biomarker analysis publication reported by Motzer et al..^49^ A total of 59 signatures were using including tumor microenvironment-derived signatures (e.g., T-cells, B-cells, Macrophages) and pathway-derived signatures (e.g., cell cycle, lipid metabolism, cell-cell signaling) and PD-L1 status (Table S8). In total 726 patients (372 sunitinib, 354 avelumab+axitinib) were retrieved. The data was split into training and testing with a 50% prevalence (random seed = 0) stratified by treatment and survival event. The PBMF was trained to identify a sub-population predictive of avelumab+axitinib against sunitinib using progressive free survival time and event as endpoints.

POSEIDON is a phase 3 randomized clinical trial that evaluated the efficacy of durvalumab plus tremelimumab plus chemotherapy and durvalumab plus chemotherapy against chemotherapy alone in first-line metastatic non-small-cell lung cancer (mNSCLC).^50^ In this study, we focused on peripheral blood RNA seq data for durvalumab + chemotherapy (114 patients) and chemotherapy alone (114 patients) treatment arms. RNA seq data was Log2(TPM+0.001) transformed, and we extracted a set of custom and publicly available tumor microenvironment-related signatures^51^ (Table S9) using the median score across genes. Dataset is split into training / testing with a 50% prevalence (random seed = 0) stratified by treatment and event. PBMF was trained using to identify predictive biomarker of durvalumab + chemotherapy against chemotherapy alone using overall survival time and event as endpoints.

Data from the Tempus NSCLC cohort were selected from the Tempus deidentified multimodal database.^52^ Patients were included if they were diagnosed with a primary or metastatic NSCLC diagnosis on or after 2016, confirmed by histology, and received chemotherapy or ICIs as first treatment. For these patients, real-world overall survival was calculated using treatment start date as the index date. RNA expression (batch-corrected and transformed to transcripts per million) data was obtained for pre-treatment samples. In the case of patients with multiple biopsies, only the closest one to treatment start date was selected. ssGSEA (corto R package) was run per RNA sample for the 50 cancer hallmark gene sets (msigDB C5).^53,54^ A total of 201 patients with stage 4 NSCLC undergoing chemotherapy (84) or immunotherapy (117) were selected. The data set was equally split into training and testing (50% each) and stratified by treatment (random seed = 0). The training set had 42 patients with chemotherapy and 58 with immunooncology treatment; and the testing set had 42 patients with chemotherapy and 59 with immunooncology treatment. We used overall survival and death as endpoints for training the PBMF model.

The POPLAR and OAK clinical trials were used to represent phases 2 and 3, respectively, to evaluate the efficacy of atezolizumab as a second-line therapy for patients unresponsive to first-line platinum-based chemotherapy in the NSCLC population. The therapeutic potential of atezolizumab was compared against that of docetaxel. The dataset, sourced from Gandara et al.,^27^ encompasses ctDNA from blood samples in addition to patient demographics and clinical biomarkers, as detailed in Table S6. We conducted a prevalence-based ranking of ctDNA genes from patients in the POPLAR trial, identifying the top 20 genes that exhibit a minimum prevalence of 20% across the combined data set from both atezolizumab and docetaxel cohorts. The PBMF was not trained by using progression-free survival, and this outcome was used for testing only. POPLAR trial data were used for training the PBMF, and OAK was used for independent evaluation. We used the overall survival time and event as endpoints. The PBMF ensemble model performance is depicted in Fig. 5c.

The CheckMate prospective clinical trials 009, 010, and 025 were designed to evaluate the efficacy of nivolumab (PD-1 blockade) against everolimus (mTOR inhibition) in advanced clear cell renal carcinoma (ccRCC). RNA sequencing (RNA-seq) and whole-exome sequencing (WES) derived features were obtained from Braun et al.^55^ The PBMF was trained using the phase 2 CheckMate 010 clinical trial data and validated on the combined populations of CheckMate 025 and CheckMate 009. We included only patients with a complete set of features, excluding any with missing data. Consequently, 199 patients out of the available 311 had all complete features. Among these, 25 patients were from the Phase 2 (CheckMate 010) clinical trial. As CheckMate 010 did not have a control arm, we randomly selected 25 patients from the CheckMate 025 everolimus arm to match the number of patients treated with nivolumab. The remaining patients from CheckMate 009 and the Phase 3 CheckMate 025 trial were utilized for independent validation (i.e. test data set). Overall survival time and event status were used as endpoints for training the PBMF. The performance of the PBMF model on the test data set after pruning is presented in Figs. 3, 4e and the complete list of features used for training are shown in the Table S7.

IMvigor210 is a single-arm phase 2 clinical trial evaluating the efficacy of atezolizumab as a first (1L) or second (2+) line of treatment in locally advanced or metastatic urothelial carcinoma (mUC). IMvigor211 is a randomized phase 3 clinical trial that evaluated the efficacy of atezolizumab compared to chemotherapy in metastatic urothelial carcinoma as a second (2+) line of treatment. Data from IMvigor210 and 211 was downloaded from supplementary material of Yuen et al..^48^ Both studies reported a total of 1222 patients. We only kept patients without missing values and filtered out all patients that were treated with Atezo as a first line of treatment in order to match the phase 3 (IMvigor211) population. In total we obtained 691 patients (422 atezolzumab and 269 chemotherapy). For training, we selected all patients from the IMvigor210 atezolizumab arm. As control, we selected 100 patients from the chemotherapy arm from the IMvigor211 phase 3 trial. For test data, we used all the patients on the phase 3 (IMvigor211), except the patients from chemotherapy that were used during training. The features in these cohorts include: age, sex, liver metastasis, ECOG, plasma IL8 at baseline (C1D1) and after treatment IL8 (C3D1) as well as plasma IL8 ratio (C3D1/C1D1). Therefore, this analysis is not limited to baseline measurements as on-treatment increased expression of plasma IL8 are known to be predictive of worse overall survival for atezolizumab and not for chemotherapy ^48^. The PBMF was trained to identify predictive biomarkers of atezolizumab against chemotherapy using overall survival time and event in the IMvigor210 cohort and validated on the IMvigor211 trial.

### Statistical modeling and model evaluation metrics

Hazard ratios and 95% confidence intervals were computed by fitting a univariate Cox proportional hazards model (lifelines Python package) to the survival data, within a given PBMF biomarker group, and using the treatment as the only covariate. P-values for hazard ratios were computed with a Wald test. When comparing survival distributions across treatments for a given biomarker group, a logrank test statistic and its associated p-value was computed and reported. Because our analyses are all retrospective, we avoid specifying statistical significance thresholds and instead faithfully report all p-values.

Model performance on synthetic datasets was evaluated using the AUPRC metric. This was chosen because we assume that identification of biomarker positive individuals is most important for biomarker discovery, and that a minority of individuals will be biomarker positive for any given real data cohort. Therefore, metrics that equally weight model performance in identifying biomarker positives and negatives, such as area under the receiving operator characteristic curve, may be poor choices. AUPRC was not reported for clinical datasets due to lack of ground truth.

## Data Availability

All human data used are available online at the following URLs: (1) https://www.uniklinik-freiburg.de/imbi/stud-le/multivariable-model-building.html (2) https://www.nature.com/articles/s41591-018-0134-3. Simulation data are available upon reasonable request to authors.

https://www.nature.com/articles/s41591-018-0134-3

## Data availability

Data for breast cancer cohorts is available at the following URL: https://www.uniklinik-freiburg.de/imbi/stud-le/multivariable-model-building.html. Data for diabetic retinopathy cohort is available within the R survival package.^17^ Data for POPLAR and OAK studies was accessed from Gandara et al.^27^ Data from Tempus may be purchased for use (https://www.tempus.com). IMmotion150, IMVigor210 and IMVigor211 data can be obtained directly from Yuen et al.^48^ supplementary material. CheckMate data can be downloaded from the supplementary information from Braun et al.^55^ publication. JAVELIN 101 Renal can be obtained directly from Motzer et al..^49^ publication. POSEIDON data underlying the findings described in this manuscript may be obtained in accordance with AstraZeneca’s data sharing policy described at https://astrazenecagrouptrials.pharmacm.com/ST/Submission/Disclosure. Data for studies directly listed on Vivli can be requested through Vivli at www.vivli.org. Data for studies not listed on Vivli could be requested through Vivli at https://vivli.org/members/enquiries-about-studies-not-listed-on-the-vivli-platform/. The AstraZeneca Vivli member page is also available outlining further details: https://vivli.org/ourmember/astrazeneca/. Requests to access these datasets should be directed to www.vivli.org.

## Code availability

Code for the PBMF and to reproduce the analyses and simulations in this manuscript will be made publicly available on Github.

## Acknowledgments

We thank J.C. Barrett and A. Meier for discussions of this work. We thank D.J. Shuman for editing help.

## Author contributions

G.A.-A. contributed to the conception of the study. G.A-A., D.E.B., G.J.S., E.K. and E.J. contributed to the design of the study. G.A-A., D.E.B., G.J.S., E.K., K.M.S., and E.J. contributed to algorithm development. G.A-A., D.E.B., G.J.S., K.M.S., and S.C.P. contributed to analysis of the data. G.A-A., D.E.B., G.J.S., and E.J. wrote the manuscript. E.J. supervised the work.

## Additional information

Supplementary Information is available for this paper.

Correspondence and requests for materials should be addressed to Gustavo Arango-Argoty and Etai Jacob.

