## Supplementary material for "AI-Driven Predictive Biomarker Discovery with Contrastive Learning to Improve Clinical Trial Outcomes": SI

**Supplementary Information**

### Supplementary methods

#### Creating uncorrelated covariate matrix for simulations

To properly control the relationship between covariates, we simulated random multivariate normal with specific covariance matrix. Although induced covariance may be interesting in some cases, we forced all the features to be fully uncorrelated by using an identity matrix as a covariance matrix. To ease the definitions of the hazard ratio group in the next stage, a binarization process was performed in each feature. To create a particular prevalence, the binarization was done at the specific percentile desired in each feature. This step was followed by multiplication of the features and treatment to the level needed (i.e., first-order interaction, two feature interactions or higher). The next step is the definition of the underlying hazard ratios structure, meaning the definition of how many different groups will be in the data and the relative hazard between them. Once the values of the parametric model are defined, the covariate extended matrix is used to create survival times by sampling from the inverted hazard function. Once the times are obtained, it is possible to increase the complexity of the data set by inducing extra noise in several ways. For example, while fully random features can be added at the beginning in the covariate matrix and then have their parameters in the hazard function set to 1, extra noise can be added to the predictive and prognostic features to transform the features from binary back to continuous.

### Optimizing Virtual Twins for simulations

To perform a fair comparison between the performance of virtual twins and the PBMF, a grid search was conducted over various hyperparameters for the random forest used in the virtual twins model. Different values for the number of features chosen in each split, the maximum depth of each tree, and the number of trees in the forest were evaluated (Table S5). Similar data to the one used during the experiments was created. In this case a dataset containing the predictive biomarker (2 features) in addition to 3 random features was used to train and evaluate these models. For each combination of hyperparameters, 10 different splits of the training and test set were generated from 80% of the data. After evaluating the results for each model trained on a different split of the data, we recorded the mean and standard deviation of the AUPRC for both the training and test sets. The best-performing model from the search was trained with 4 features in each split, 200 trees in the ensemble, and a maximum depth of 5, which resulted in a training AUPRC of 0.946 ± 0.014 and a testing AUPRC of 0.933 ± 0.015.

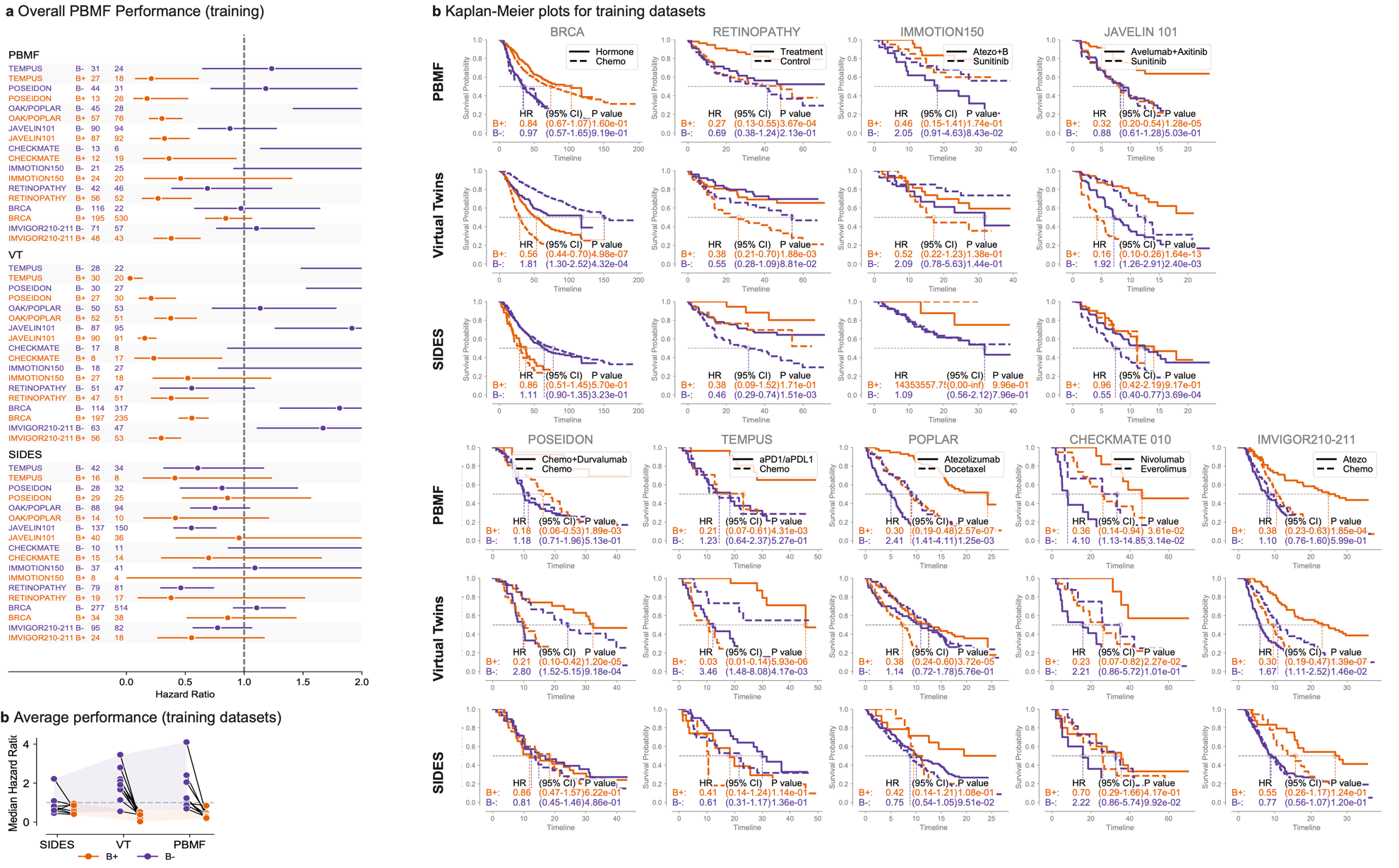

**Fig. S1.** Training performance of the predictive biomarker modeling framework (PBMF) ensemble after pruning.

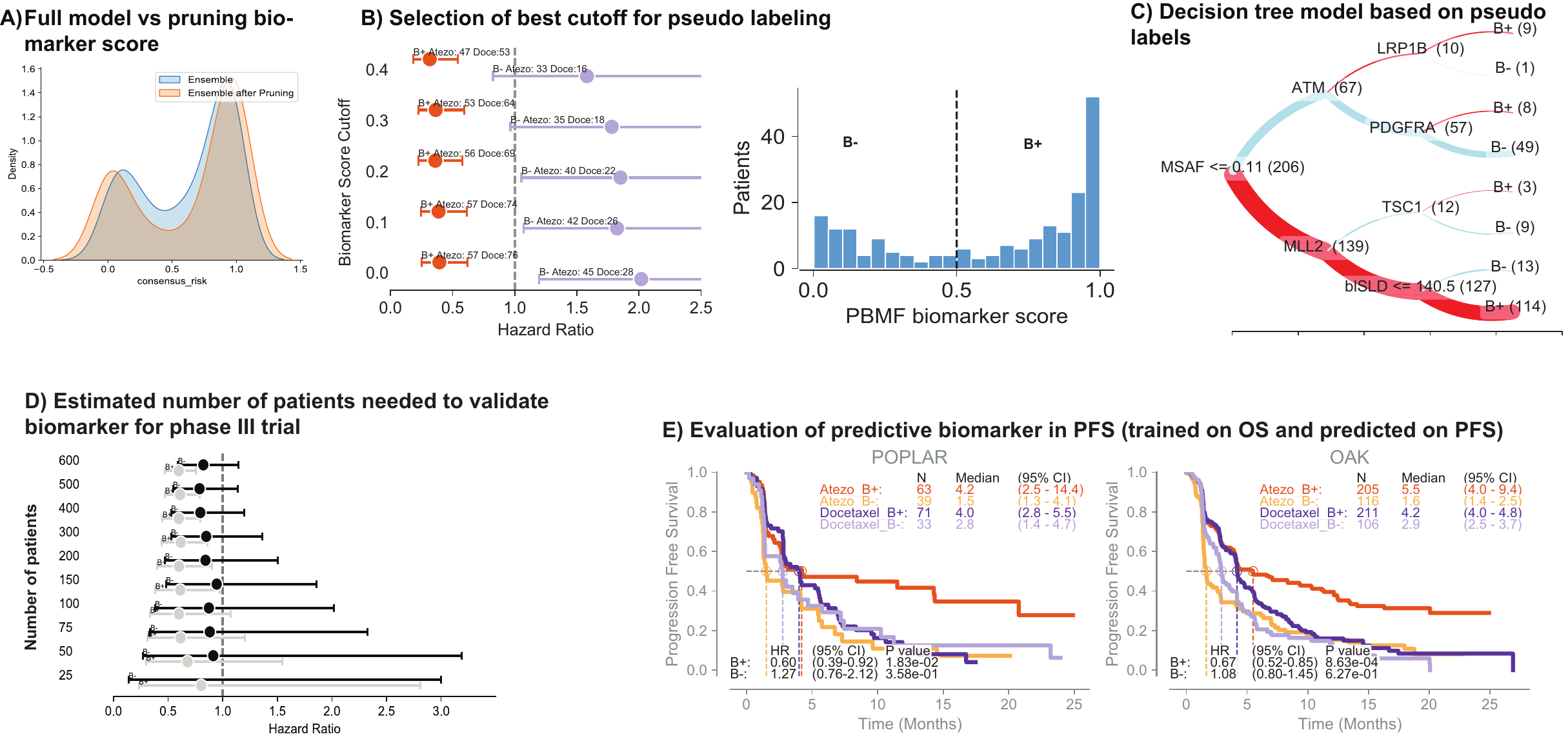

**Figure S2. a**, Distribution of ensemble versus pruned model scores. **b**, Using the pruned model, identify the best cutoff to achieve high-confidence biomarker pseudo-labels. **c**, Decision tree depicting a set of rules that simplifies the predictive biomarker. Red lines, B+; blue lines, B–; line thickness is proportional to number of patients in parenthesis. **d**, Estimation of the minimum number of patients required to validate a predictive biomarker discovered in a phase 2 (POPLAR) clinical trial. Data were subsampled from the OAK trial. **e**, Evaluation of consistency across endpoints. PBMF was trained on overall survival (OS) endpoint and evaluated by using the tree-derived predictive biomarker on progression-free survival (PFS).

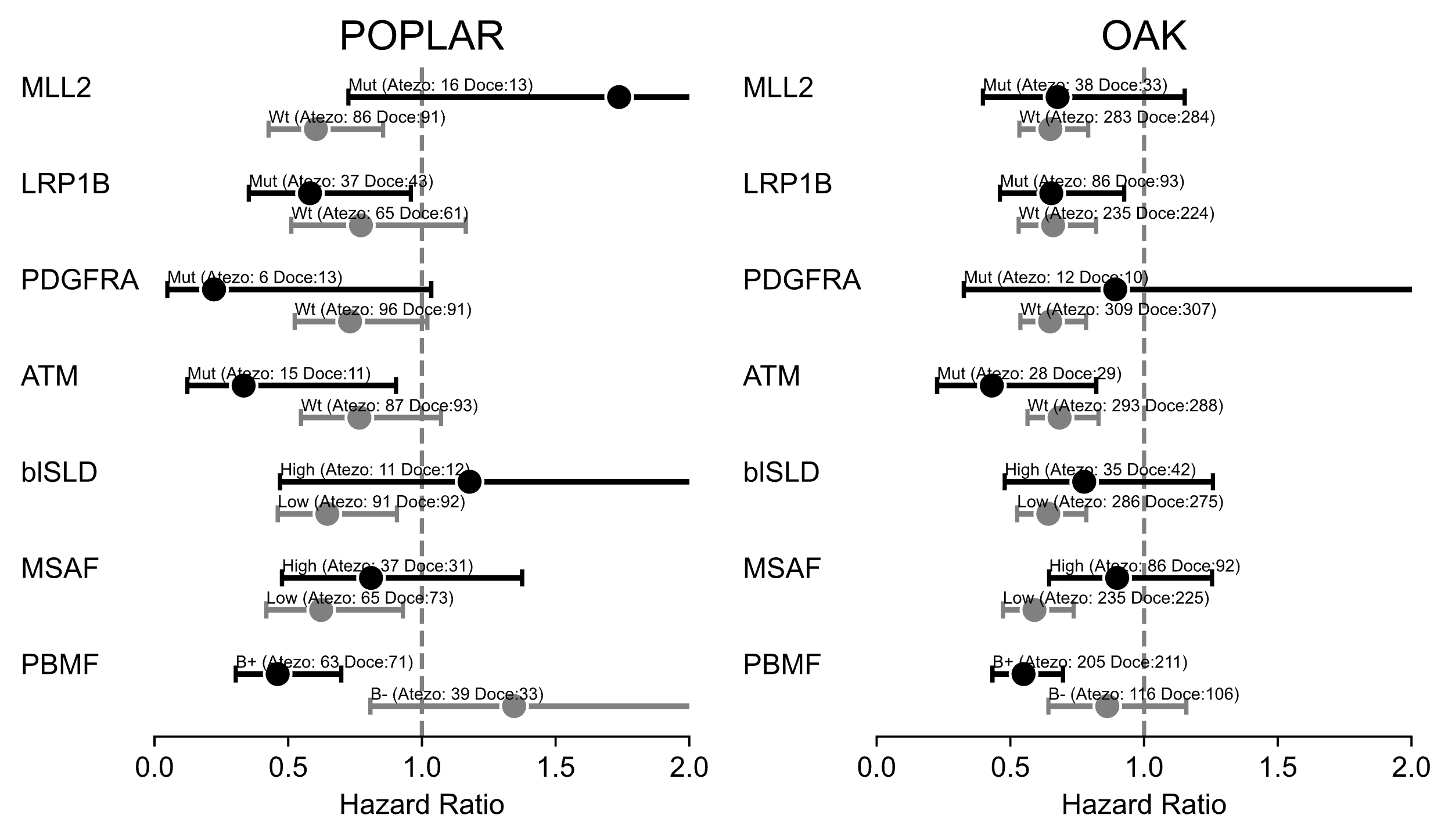

**Figure S3** Forest plot depicting the predictive value of individual tree selected features. Cutoffs for continuous features are taken from the tree classifier. Mutational status was defined as wild type (*wt*) or mutated (*mut*).

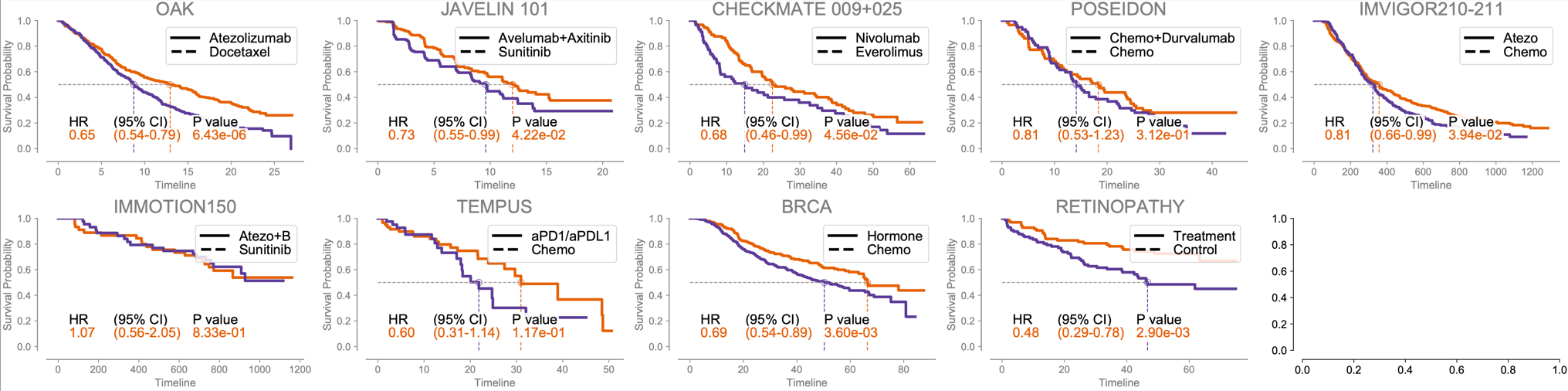

**Figure S4** Kaplan-Meier curves colored by treatment for the biomarker-evaluable population for the test sets for each clinical study evaluated. Note that in the case where the study was split into train/test cohorts, these will not be the same biomarker-evaluable populations reported for that given study.

**Table S1.** Model hyperparameters for identifying predictive biomarkers for clinical studies

* Value varied depending on the simulation experiment run.

| PBMF  parameters | **Rotterdam and German breast cancer studies** | **Tempus Non-small cell lung cancer** | **POPLAR and OAK** | **Checkmate 009-010-025** | **Immotion 150** | **Imvigor 210 - 211** | **JAVELIN** | **POSEIDON** | **RETINOPATHY** | **Simulations** |
| --- | --- | --- | --- | --- | --- | --- | --- | --- | --- | --- |
| Seed | 0 | 0 | 0 | 0 | 0 | 0 | 0 | 0 | 0 | 0 |
| Number of models | 100 | 500 | 100 | 100 | 100 | 100 | 100 | 100 | 100 | 128/1024 |
| Subset training data | 80% | 80% | 80% | 80% | 95% | 90% | 90% | 90% | 90% | 80% |
| Ignore patients during loss computation | 10% | 10% | 10% | 10% | 5% | 10% | 10% | 10% | 10% | 10% |
| Use only *n* features (for each model if using an ensemble) | 4 of 7 | 30 of 50 | 28 of 29 | 37 of 38 | 8 of 8 | 6 of 7 | 58 of 59 | 34 of 35 | 3 of 3 | * |
| Learning rate | 0.01 | 0.01 | 0.01 | 0.01 | 0.01 | 0.01 | 0.01 | 0.01 | 0.01 | 0.01 |
| Layers [neurons] | [64] | [64] | [64] | [64] | [64] | [64] | [64] | [64] | [64] | [64] |
| Training epochs | 500 | 100 | 500 | 100 | 100 | 1000 | 100 | 50 | 200 | 2000 |
| Minimum population | 0.75 | 0.5 | NA | NA | 0.5 | 0.5 | 0.5 | NA | 0.5 | 0.5 |
| w1, w2 | 1, 1 | 1, 1 | 1, 0** | 1,0** | 1, 1 | 1,1 | 1,1 | 1,0** | 1, 1 | 1, 1 |
| L1, L2 normalization | 0, 0 | 0, 0 | 0, 0 | 0,0 | 0, 0 | 0,0 | 0,0 | 0,0 | 0, 0 | 0, 0 |
| pruning percentile | 95 | 95 | 95 | 95 | 90 | 90 | 95 | 90 | 90 | 90 |

** Prevalence loss deactivated.

| Virtual twins param-eters | **Rotterdam and German breast cancer studies** | **Tempus Non-small cell lung cancer** | **POPLAR and OAK** | **Checkmate 009-010-025** | **Immotion 150** | **Imvigor 210 - 211** | **JAVELIN** | **POSEIDON** | **RETINOPATHY** | **Simulations** |
| --- | --- | --- | --- | --- | --- | --- | --- | --- | --- | --- |
| Seed | 1 | 1 | 1 | 1 | 1 | 1 | 1 | 1 | 1 | 1 |
| Max features | sqrt | Sqrt | Sqrt | Sqrt | Sqrt | Sqrt | Sqrt | Sqrt | Sqrt | sqrt |
| Max_depth | 5 | 5 | 5 | 5 | 5 | 5 | 5 | 5 | 5 | 5 |
| Min node size | 10 | 10 | 10 | 5 | 10 | 10 | 10 | 10 | 20 | 10 |
| Num trees | 100 | 100 | 100 | 100 | 100 | 100 | 100 | 100 | 500 | 100 |

| **SIDES param-eters** | **Rotterdam and German breast cancer studies** | **Tempus Non-small cell lung cancer** | **POPLAR and OAK** | **Checkmate 009-010-025** | **Immotion 150** | **Imvigor 210 - 211** | **JAVELIN** | **POSEIDON** | **RETINOPATHY** |
| --- | --- | --- | --- | --- | --- | --- | --- | --- | --- |
| Min subgroup size | 30 | 10 | 10 | 10 | 10 | 10 | 10 | 10 | 30 |
| Criterion tpe | 1 | 1 | 1 | 1 | 1 | 1 | 1 | 1 | 1 |
| Depth | 3 | 3 | 3 | 3 | 3 | 3 | 3 | 3 | 3 |
| Width | 5 | 5 | 5 | 5 | 5 | 5 | 5 | 5 | 5 |
| Gamma | NA | NA | NA | NA | NA | NA | NA | NA | NA |
| Local mult adj | 1 | 1 | 1 | 1 | 1 | 1 | 1 | 1 | 1 |
| N perms mult adjust | 200 | 200 | 200 | 200 | 200 | 200 | 200 | 200 | 200 |
| algorithm | Adaptive SIDES | SIDES | SIDES | SIDES | SIDES | SIDES | SIDES | SIDES | Adaptive SIDES |
| Top biomarkers | 2 | 2 | 2 | 2 | 2 | 2 | 2 | 2 | 2 |

**Table S2.** AUPRC on the test set for different training size comparing VT and PBMF^a^

| **Size** | **VT (AUPRC)** | **PBMF (AUPRC)** | **PBMFp75 (AUPRC)** | **PBMFp90 (AUPRC)** |
| --- | --- | --- | --- | --- |
| 250 | 0.752 ± 0.091 | 0.786 ± 0.066 | 0.785 ± 0.070 | 0.787 ± 0.068 |
| 500 | 0.803 ± 0.055 | 0.849 ± 0.065 | 0.844 ± 0.074 | 0.845 ± 0.073 |
| 1000 | 0.858 ± 0.029 | 0.918 ± 0.047 | 0.918 ± 0.060 | 0.917 ± 0.059 |
| 2000 | 0.858 ± 0.030 | 0.966 ± 0.028 | 0.964 ± 0.044 | 0.963 ± 0.045 |
| 4000 | 0.845 ± 0.026 | 0.985 ± 0.019 | 0.981 ± 0.038 | 0.980 ± 0.041 |

AUPRC, area under the precision-recall curve; VT, virtual twins.

^a^ A total of 128 ensembles, 3 features from a data set containing 3 features. p75 and p90 are the corresponding models pruned to the top 25 and top 10 models in the ensemble.

**Table S3.** Patient stratification using most important tree-derived features^a^

| **HR** | **HR_lo** | **HR_hi** | ***P*** | **Atezo (*N*)** | | **Doce (*N*)** | | **Data set** | **Biomarker** | **Group** |
| --- | --- | --- | --- | --- | --- | --- | --- | --- | --- | --- |
| 0.460543 | 0.303604 | 0.698606 | 2.65E-04 | 63 |  | 71 |  | POPLAR | PBMF | B+ |
| 1.344858 | 0.807394 | 2.240101 | 2.55E-01 | 39 |  | 33 |  | POPLAR | PBMF | B– |
| 0.549327 | 0.432999 | 0.696908 | 8.05E-07 | 205 |  | 211 |  | OAK | PBMF | B+ |
| 0.862457 | 0.642394 | 1.157906 | 3.25E-01 | 116 |  | 106 |  | OAK | PBMF | B– |
| 0.809564 | 0.47673 | 1.374771 | 4.34E-01 | 37 |  | 31 |  | POPLAR | MSAF | High |
| 0.62323 | 0.4182 | 0.928778 | 2.02E-02 | 65 |  | 73 |  | POPLAR | MSAF | Low |
| 0.899465 | 0.64495 | 1.254421 | 5.32E-01 | 86 |  | 92 |  | OAK | MSAF | High |
| 0.590117 | 0.47252 | 0.73698 | 3.30E-06 | 235 |  | 225 |  | OAK | MSAF | Low |
| 1.178311 | 0.469664 | 2.956195 | 7.27E-01 | 11 |  | 12 |  | POPLAR | blSLD | High |
| 0.64637 | 0.461001 | 0.906275 | 1.14E-02 | 91 |  | 92 |  | POPLAR | blSLD | Low |
| 0.776102 | 0.478992 | 1.257506 | 3.03E-01 | 35 |  | 42 |  | OAK | blSLD | High |
| 0.641929 | 0.52561 | 0.783989 | 1.39E-05 | 286 |  | 275 |  | OAK | blSLD | Low |
| 1.737477 | 0.725244 | 4.162495 | 2.15E-01 | 16 |  | 13 |  | POPLAR | MLL2 | Mut |
| 0.603687 | 0.42629 | 0.854906 | 4.47E-03 | 86 |  | 91 |  | POPLAR | MLL2 | Wt |
| 0.676654 | 0.397474 | 1.151929 | 1.50E-01 | 38 |  | 33 |  | OAK | MLL2 | Mut |
| 0.649714 | 0.533546 | 0.791174 | 1.78E-05 | 283 |  | 284 |  | OAK | MLL2 | Wt |
| 0.333364 | 0.123031 | 0.90328 | 3.08E-02 | 15 |  | 11 |  | POPLAR | ATM | Mut |
| 0.766136 | 0.547877 | 1.071342 | 1.19E-01 | 87 |  | 93 |  | POPLAR | ATM | Wt |
| 0.430885 | 0.226066 | 0.821272 | 1.05E-02 | 28 |  | 29 |  | OAK | ATM | Mut |
| 0.684175 | 0.564204 | 0.829655 | 1.14E-04 | 293 |  | 288 |  | OAK | ATM | Wt |
| 0.581416 | 0.352588 | 0.958752 | 3.36E-02 | 37 |  | 43 |  | POPLAR | LRP1B | Mut |
| 0.771924 | 0.511864 | 1.164113 | 2.17E-01 | 65 |  | 61 |  | POPLAR | LRP1B | Wt |
| 0.653204 | 0.460844 | 0.925856 | 1.67E-02 | 86 |  | 93 |  | OAK | LRP1B | Mut |
| 0.660094 | 0.530953 | 0.820647 | 1.84E-04 | 235 |  | 224 |  | OAK | LRP1B | Wt |
| 0.222601 | 0.04788 | 1.0349 | 5.53E-02 | 6 |  | 13 |  | POPLAR | PDGFRA | Mut |
| 0.731323 | 0.524079 | 1.020521 | 6.57E-02 | 96 |  | 91 |  | POPLAR | PDGFRA | Wt |
| 0.892299 | 0.326007 | 2.44227 | 8.24E-01 | 12 |  | 10 |  | OAK | PDGFRA | Mut |
| 0.648723 | 0.537572 | 0.782855 | 6.39E-06 | 309 |  | 307 |  | OAK | PDGFRA | Wt |

Atezo, atezolizumab; Doce, docetaxel.

^a^Endpoint for all data sets was overall survival.

**Table S4.** Efficacy comparison of PBMF against blood TMB (16 bp/Mb cutoff)

| **HR** | **HR_lo** | **HR_hi** | ***P*** | **Atezo (*N*)** | **Doce (*N*)** | **Data set** | **Endpoint** | **Biomarker** | **Group** |
| --- | --- | --- | --- | --- | --- | --- | --- | --- | --- |
| 0.460543 | 0.303604 | 0.698606 | 2.65E-04 | 63 | 71 | POPLAR | OS | PBMF | B+ |
| 0.737623 | 0.509153 | 1.068614 | 1.08E-01 | 63 | 71 | POPLAR | PFS | PBMF | B+ |
| 1.344858 | 0.807394 | 2.240101 | 2.55E-01 | 39 | 33 | POPLAR | OS | PBMF | B– |
| 1.335188 | 0.813648 | 2.19103 | 2.53E-01 | 39 | 33 | POPLAR | PFS | PBMF | B– |
| 0.549327 | 0.432999 | 0.696908 | 8.05E-07 | 205 | 211 | OAK | OS | PBMF | B+ |
| 0.787426 | 0.640296 | 0.968365 | 2.35E-02 | 205 | 211 | OAK | PFS | PBMF | B+ |
| 0.862457 | 0.642394 | 1.157906 | 3.25E-01 | 116 | 106 | OAK | OS | PBMF | B– |
| 1.11819 | 0.849369 | 1.472092 | 4.26E-01 | 116 | 106 | OAK | PFS | PBMF | B– |
| 0.55635 | 0.311286 | 0.994344 | 4.78E-02 | 25 | 38 | POPLAR | OS | bTMB | High |
| 0.572667 | 0.330899 | 0.991082 | 4.64E-02 | 25 | 38 | POPLAR | PFS | bTMB | High |
| 0.766408 | 0.521741 | 1.12581 | 1.75E-01 | 77 | 66 | POPLAR | OS | bTMB | Low |
| 1.117139 | 0.78627 | 1.587241 | 5.36E-01 | 77 | 66 | POPLAR | PFS | bTMB | Low |
| 0.622256 | 0.429643 | 0.901219 | 1.21E-02 | 78 | 82 | OAK | OS | bTMB | High |
| 0.657654 | 0.469874 | 0.920479 | 1.46E-02 | 78 | 82 | OAK | PFS | bTMB | High |
| 0.671064 | 0.542226 | 0.830514 | 2.45E-04 | 243 | 235 | OAK | OS | bTMB | Low |
| 0.988343 | 0.817333 | 1.195133 | 9.04E-01 | 243 | 235 | OAK | PFS | bTMB | Low |

Atezo, atezolizumab; Doce, docetaxel; OS, overall survival; PFS, progression-free survival; bTMB, blood tumor mutation burden.

**Table S5.** Results of grid search for the optimum hyperparameters for the virtual twins model for simulation benchmarks. Hyperparameters that gave the best performance in boldface.

| **No. of features** | **No. of trees** | **Maximum depth** | **AUPRC** | |
| --- | --- | --- | --- | --- |
|  |  |  | **Training set** | **Testing set** |
| 2 | 10 | 3 | 0.842 ± 0.034 | 0.819 ± 0.039 |
| 2 | 10 | 5 | 0.880 ± 0.024 | 0.857 ± 0.026 |
| 2 | 10 | 7 | 0.870 ± 0.031 | 0.840 ± 0.035 |
| 2 | 50 | 3 | 0.896 ± 0.030 | 0.882 ± 0.030 |
| 2 | 50 | 5 | 0.922 ± 0.021 | 0.907 ± 0.018 |
| 2 | 50 | 7 | 0.920 ± 0.019 | 0.904 ± 0.017 |
| 2 | 100 | 3 | 0.910 ± 0.023 | 0.894 ± 0.022 |
| 2 | 100 | 5 | 0.932 ± 0.017 | 0.914 ± 0.015 |
| 2 | 100 | 7 | 0.929 ± 0.018 | 0.911 ± 0.014 |
| 2 | 200 | 3 | 0.913 ± 0.024 | 0.897 ± 0.022 |
| 2 | 200 | 5 | 0.936 ± 0.017 | 0.918 ± 0.016 |
| 2 | 200 | 7 | 0.932 ± 0.017 | 0.915 ± 0.014 |
| 4 | 10 | 3 | 0.898 ± 0.025 | 0.881 ± 0.028 |
| 4 | 10 | 5 | 0.909 ± 0.022 | 0.893 ± 0.023 |
| 4 | 10 | 7 | 0.899 ± 0.024 | 0.884 ± 0.027 |
| 4 | 50 | 3 | 0.921 ± 0.019 | 0.908 ± 0.016 |
| 4 | 50 | 5 | 0.938 ± 0.012 | 0.928 ± 0.013 |
| 4 | 50 | 7 | 0.933 ± 0.014 | 0.923 ± 0.017 |
| 4 | 100 | 3 | 0.929 ± 0.020 | 0.917 ± 0.019 |
| 4 | 100 | 5 | 0.942 ± 0.013 | 0.932 ± 0.013 |
| 4 | 100 | 7 | 0.938 ± 0.014 | 0.927 ± 0.015 |
| 4 | 200 | 3 | 0.934 ± 0.02 | 0.92 ± 0.019 |
| **4** | **200** | **5** | **0.946 ± 0.014** | **0.933 ± 0.015** |
| 4 | 200 | 7 | 0.94 ± 0.015 | 0.927 ± 0.016 |
| 5 | 10 | 3 | 0.891 ± 0.030 | 0.873 ± 0.027 |
| 5 | 10 | 5 | 0.908 ± 0.023 | 0.89 ± 0.023 |
| 5 | 10 | 7 | 0.899 ± 0.024 | 0.878 ± 0.027 |
| 5 | 50 | 3 | 0.923 ± 0.019 | 0.912 ± 0.019 |
| 5 | 50 | 5 | 0.938 ± 0.014 | 0.929 ± 0.016 |
| 5 | 50 | 7 | 0.933 ± 0.017 | 0.923 ± 0.018 |
| 5 | 100 | 3 | 0.929 ± 0.020 | 0.918 ± 0.020 |
| 5 | 100 | 5 | 0.942 ± 0.015 | 0.931 ± 0.016 |
| 5 | 100 | 7 | 0.937 ± 0.016 | 0.926 ± 0.017 |
| 5 | 200 | 3 | 0.934 ± 0.022 | 0.921 ± 0.022 |
| 5 | 200 | 5 | 0.944 ± 0.015 | 0.932 ± 0.018 |
| 5 | 200 | 7 | 0.939 ± 0.016 | 0.926 ± 0.019 |

**Table S6** Clinical, genomic, and demographic features in the OAK and POPLAR studies

| **Feature name** | **Description** |
| --- | --- |
| BAGE | Baseline age |
| blSLD | Sum of longest diameter of target lesions at baseline |
| HIST | Histology |
| METSITES | Number of metastatic sites at enrollment |
| TOBHX | Smoking history |
| ECOGGR | ECOG group |
| MSAF | Maximum somatic allele frequency |
| bTMB | Blood TMB |
| SEX | Patient sex |
| Top 20 prevalent genes | *TP53*, *LRP1B*, *DNMT3A*, *SPTA1*, *NF1*, *FAT3*, *KEAP1*, *STAG2*, *MLL2*, *MLL3*, *ATM*, *TSC1*, *FAT1*, *EGFR*, *STK11*, *NFE2L2*, *PREX2*, *EPHA3*, *CHEK2*, *PDGFRA* |

ECOG, Eastern Cooperative Oncology Group; TMB, tumor mutation burden.

**Table S7** Clinical, genomic, and demographic features in the CheckMate 009-010-025 studies

| **Feature name** | **Description** |
| --- | --- |
| RNA signatures | 't_cells_cd8',  'leukocyte_total',  't_cells_cd4_memory_resting', |
| Molecular (CNV) | 'Amplification_1q21.3',  'Amplification_5p15.33',  'Amplification_5q31.3',  'Amplification_5q35.3',  'Amplification_7p22.2',  'Amplification_7q36.2',  'Amplification_8q24.3',  'Amplification_12q24.32',  'Amplification_17q25.3',  'Amplification_20q13.33',  'Amplification_21q22.3',  'Deletion_1p36.31',  'Deletion_1p36.11',  'Deletion_3p21.1',  'Deletion_6p22.2',  'Deletion_6p21.32',  'Deletion_6q25.2',  'Deletion_9p21.3',  'Deletion_9q34.3',  'Deletion_10q23.31',  'Deletion_10q26.3',  'Deletion_14q32.33',  'Deletion_19p13.3',  'Deletion_19q13.42' |
| Molecular (Mutations) | 'VHL',  'PBRM1',  'SETD2',  'BAP1',  'KDM5C',  'MTOR',  'TP53',  'PTEN',  'TSC1',  'NF2',  'PIK3CA' |

**Table S8** Features used in the JAVELIN 101 Renal study

| **Feature name** | **Description** |
| --- | --- |
| PDL1 | Pdl1_status |
| RNA expression signatures (TME + pathways) | 'B9991003_rc9_Cell_cycle',  'LM22.Eosinophils',  'B9991003_c8_Immune_response',  'B9991003_c9_Cell_cycle',  'LM22.T_cells_regulatory_Tregs',  'B9991003_rc1_Lipid_metabolic_process',  'B9991003_rc5_Cell_cell_signaling',  'LM22.Macrophages_M2',  'LM22.Macrophages_M0',  'LM22.Macrophages_M1',  'LM22.NK_cells_resting',  'B9991003_c14_Oxygen_transport',  'B9991003_c11_EMT',  'MCDERMOTT_ANGIO',  'MCDERMOTT_TEFF',  'B9991003_c2_Organic_acid_metabolic_process',  'B9991003_c16_Neutrophils',  'LM22.T_cells_gamma_delta',  'LM22.B_cells_naive',  'B9991003_c6_Negative_regulation_of_EMT',  'LM22.T_cells_CD4_memory_resting',  'B9991003_c7_Transmembrane_transport',  'B9991003_rc2_Organic_acid_metabolic_process',  'MCDERMOTT_MYELOID_INFLAMMATION',  'B9991003_c1_Lipid_metabolic_process',  'B9991003_c18_B_cell_activation',  'DUX4_TF_signature',  'B9991003_c22_Morphogenesis_Homeobox_gene_c',  'B9991003_c20_Interferon_gamma_response',  'LM22.Mast_cells_resting',  'LM22.Dendritic_cells_activated',  'Tcell_inflammed_GEP_MERCK',  'B9991003_c21_Chromosome_Y_linked',  'B9991003_c23_Cell_adhesion_at_cell_membrane',  'LM22.T_cells_CD4_naive',  'LM22.B_cells_memory',  'B9991003_c3_Angiogenesis',  'B9991003_c5_Cell_cell_signaling',  'B9991003_c15_Skin_development',  'B9991003_c19_Plasma_cells',  'B9991003_rc11_EMT',  'LM22.Dendritic_cells_resting',  'B9991003_Javelin_Renal_101_genes26',  'B9991003_rc13_Cytoskeleton_organization',  'B9991003_c10_Myogenesis',  'B9991003_rc3_Angiogenesis',  'B9991003_c13_Cytoskeleton_organization',  'B9991003_c4_Cell_development',  'LM22.Monocytes',  'LM22.T_cells_follicular_helper',  'LM22.NK_cells_activated',  'LM22.T_cells_CD4_memory_activated',  'B9991003_c12_TNFa_signaling_via_NFkB',  'LM22.Mast_cells_activated',  'LM22.T_cells_CD8',  'LM22.Plasma_cells',  'LM22.Neutrophils',  'B9991003_c17_Glucocorticoid_metabolic_process' |

**Table S9** RNA expression signatures used for peripheral blood analysis in POSEIDON

| Angiogenesis | VEGFA, VEGFB, VEGFC, PDGFC, CXCL8, CXCR2, FLT1, PGF, CXCL5, KDR, ANGPT1, ANGPT2, TEK, VWF, CDH5 | ENSG00000112715, ENSG00000173511, ENSG00000150630, ENSG00000145431, ENSG00000169429, ENSG00000180871, ENSG00000102755, ENSG00000119630, ENSG00000163735, ENSG00000128052, ENSG00000154188, ENSG00000091879, ENSG00000120156, ENSG00000110799, ENSG00000179776 |
| --- | --- | --- |
| Antitumor cytokines | TNF, IFNB1, IFNA2, CCL3, TNFSF10, IL21 | ENSG00000232810, ENSG00000171855, ENSG00000188379, ENSG00000277632, ENSG00000121858, ENSG00000138684 |
| B_cell | MS4A1, CD19, CD22, CD79A | ENSG00000156738, ENSG00000177455, ENSG00000012124, ENSG00000105369 |
| CD1c | CD1C, CLEC10A, CD1E | ENSG00000158481, ENSG00000132514, ENSG00000158488 |
| Cancer-associated fibroblasts | COL1A1, COL1A2, COL5A1, ACTA2, FGF2, FAP, LRP1, CD248, COL6A1, COL6A2, COL6A3, CXCL12, FBLN1, LUM, MFAP5, MMP3, MMP2, PDGFRB, PDGFRA | ENSG00000108821, ENSG00000164692, ENSG00000130635, ENSG00000107796, ENSG00000138685, ENSG00000078098, ENSG00000123384, ENSG00000174807, ENSG00000142156, ENSG00000142173, ENSG00000163359, ENSG00000107562, ENSG00000077942, ENSG00000139329, ENSG00000197614, ENSG00000149968, ENSG00000087245, ENSG00000113721, ENSG00000134853 |
| Checkpoint molecules | PDCD1, CD274, CTLA4, LAG3, PDCD1LG2, BTLA, HAVCR2, TIGIT, VSIR | ENSG00000188389, ENSG00000120217, ENSG00000163599, ENSG00000089692, ENSG00000197646, ENSG00000186265, ENSG00000135077, ENSG00000181847, ENSG00000107738 |
| Co-activation molecules | CD28, CD40, TNFRSF4, ICOS, TNFRSF9, CD27, CD80, CD86, CD40LG, CD83, TNFSF4, ICOSLG, TNFSF9, CD70 | ENSG00000178562, ENSG00000101017, ENSG00000186827, ENSG00000163600, ENSG00000049249, ENSG00000139193, ENSG00000121594, ENSG00000114013, ENSG00000102245, ENSG00000112149, ENSG00000117586, ENSG00000160223, ENSG00000125657, ENSG00000125726 |
| EMT signature | SNAI1, SNAI2, TWIST1, TWIST2, ZEB1, ZEB2, CDH2 | ENSG00000124216, ENSG00000019549, ENSG00000122691, ENSG00000233608, ENSG00000148516, ENSG00000169554, ENSG00000170558 |
| Effector cell traffic | CXCL9, CXCL10, CXCL11, CX3CL1, CCL3, CCL4, CX3CR1, CCL5, CXCR3 | ENSG00000138755, ENSG00000169245, ENSG00000169248, ENSG00000006210, ENSG00000277632, ENSG00000275302, ENSG00000168329, ENSG00000271503, ENSG00000186810 |
| Effector cells | IFNG, GZMA, GZMB, PRF1, GZMK, ZAP70, GNLY, FASLG, TBX21, EOMES, CD8A, CD8B | ENSG00000111537, ENSG00000145649, ENSG00000100453, ENSG00000180644, ENSG00000113088, ENSG00000115085, ENSG00000115523, ENSG00000117560, ENSG00000073861, ENSG00000163508, ENSG00000153563, ENSG00000172116 |
| Endothelium | NOS3, KDR, FLT1, VCAM1, VWF, CDH5, MMRN1, ENG, CLEC14A, MMRN2 | ENSG00000164867, ENSG00000128052, ENSG00000102755, ENSG00000162692, ENSG00000110799, ENSG00000179776, ENSG00000138722, ENSG00000106991, ENSG00000176435, ENSG00000173269 |
| Granulocyte traffic | CXCL8, CXCL1, CXCL2, CXCL5, CCL11, KITLG, CXCR1, CXCR2, CCR3 | ENSG00000169429, ENSG00000163739, ENSG00000081041, ENSG00000163735, ENSG00000172156, ENSG00000049130, ENSG00000163464, ENSG00000180871, ENSG00000183625 |
| IFNG | IFNG, CD274, CXCL9, LAG3 | ENSG00000111537, ENSG00000120217, ENSG00000138755, ENSG00000089692 |
| Immune Suppression by Myeloid Cells | IDO1, ARG1, IL10, CYBB, PTGS2, IL4I1, IL6 | ENSG00000131203, ENSG00000118520, ENSG00000136634, ENSG00000165168, ENSG00000073756, ENSG00000104951, ENSG00000136244 |
| M1 signature | NOS2, TNF, IL1B, SOCS3, CMKLR1, IRF5, IL12A, IL12B, IL23A | ENSG00000007171, ENSG00000232810, ENSG00000125538, ENSG00000184557, ENSG00000174600, ENSG00000128604, ENSG00000168811, ENSG00000113302, ENSG00000110944 |
| MHC_I | B2M, TAP1, TAP2, TAPBP, NLRC5, HLA-A, HLA-B, HLA-C | ENSG00000166710, ENSG00000168394, ENSG00000204267, ENSG00000231925, ENSG00000140853, ENSG00000206503, ENSG00000234745, ENSG00000204525 |
| MHC_II | HLA-DQA1, HLA-DQB1, HLA-DRA, HLA-DRB1, HLA-DMA, HLA-DMB, HLA-DPB1 | ENSG00000196735, ENSG00000179344, ENSG00000204287, ENSG00000196126, ENSG00000204257, ENSG00000242574, ENSG00000223865 |
| Macrophage and DC traffic | CCL2, CCL7, CCL8, XCL1, CCR2, XCR1, CSF1R, CSF1 | ENSG00000108691, ENSG00000108688, ENSG00000108700, ENSG00000143184, ENSG00000121807, ENSG00000173578, ENSG00000182578, ENSG00000184371 |
| Matrix | FN1, COL1A1, COL1A2, COL4A1, COL3A1, VTN, LGALS7, LGALS9, LAMA3, LAMB3, LAMC2, TNC, ELN, COL5A1, COL11A1 | ENSG00000115414, ENSG00000108821, ENSG00000164692, ENSG00000187498, ENSG00000168542, ENSG00000109072, ENSG00000205076, ENSG00000168961, ENSG00000053747, ENSG00000196878, ENSG00000058085, ENSG00000041982, ENSG00000049540, ENSG00000130635, ENSG00000060718 |
| Matrix remodeling | CA9, MMP9, MMP2, MMP1, MMP3, MMP12, MMP7, MMP11, PLOD2, ADAMTS4, ADAMTS5, LOX | ENSG00000107159, ENSG00000100985, ENSG00000087245, ENSG00000196611, ENSG00000149968, ENSG00000262406, ENSG00000137673, ENSG00000099953, ENSG00000152952, ENSG00000158859, ENSG00000154736, ENSG00000113083 |
| Myeloid cells traffic | CSF2, CSF3, CXCL12, CCL26, IL6, CXCL8, CXCL5, CSF1R, CSF2RA, CSF3R, CXCR4, IL6R, CXCR2, CCL15, CSF1 | ENSG00000164400, ENSG00000108342, ENSG00000107562, ENSG00000006606, ENSG00000136244, ENSG00000169429, ENSG00000163735, ENSG00000182578, ENSG00000198223, ENSG00000119535, ENSG00000121966, ENSG00000160712, ENSG00000180871, ENSG00000275718, ENSG00000184371 |
| NK | GNLY, KLRC3, KLRD1, KLRF1, NCR1 | ENSG00000115523, ENSG00000205810, ENSG00000134539, ENSG00000150045, ENSG00000189430 |
| Neutrophil signature | MPO, ELANE, PRTN3, CTSG, CXCR1, CXCR2, FCGR3B, CD177, FFAR2, PGLYRP1 | ENSG00000005381, ENSG00000197561, ENSG00000196415, ENSG00000100448, ENSG00000163464, ENSG00000180871, ENSG00000162747, ENSG00000204936, ENSG00000126262, ENSG00000008438 |
| Protumor cytokines | IL10, TGFB1, TGFB2, TGFB3, IL22, MIF, IL6 | ENSG00000136634, ENSG00000105329, ENSG00000092969, ENSG00000119699, ENSG00000127318, ENSG00000240972, ENSG00000136244 |
| T cells | TBX21, ITK, CD3D, CD3E, CD3G, TRAC, TRBC1, TRBC2, CD28, CD5, TRAT1 | ENSG00000073861, ENSG00000113263, ENSG00000167286, ENSG00000198851, ENSG00000160654, ENSG00000277734, ENSG00000211751, ENSG00000211772, ENSG00000178562, ENSG00000110448, ENSG00000163519 |
| TLS_Chemokine | CCL2, CCL3, CCL4, CCL5, CCL8, CCL18, CCL19, CCL21, CXCL9, CXCL10, CXCL11, CXCL13 | ENSG00000108691, ENSG00000277632, ENSG00000275302, ENSG00000271503, ENSG00000108700, ENSG00000275385, ENSG00000172724, ENSG00000137077, ENSG00000138755, ENSG00000169245, ENSG00000169248, ENSG00000156234 |
| TLS_TFH | CXCL13, CD200, FBLN7, ICOS, SGPP2, SH2D1A, TIGIT, PDCD1 | ENSG00000156234, ENSG00000091972, ENSG00000144152, ENSG00000163600, ENSG00000163082, ENSG00000183918, ENSG00000181847, ENSG00000188389 |
| T_agonist | ICOS, CD28, CD27, TNFSF14, CD40LG, TNFRSF9, TNFRSF4, TNFRSF25, TNFRSF18, TNFRSF8, SLAMF1, CD2, CD226 | ENSG00000163600, ENSG00000178562, ENSG00000139193, ENSG00000125735, ENSG00000102245, ENSG00000049249, ENSG00000186827, ENSG00000215788, ENSG00000186891, ENSG00000120949, ENSG00000117090, ENSG00000116824, ENSG00000150637 |
| T_effector | CD8A, GZMA, GZMB, IFNG, EOMES, TBX21, CXCL9, CXCL10, KLRG1, CD44, CD37 | ENSG00000153563, ENSG00000145649, ENSG00000100453, ENSG00000111537, ENSG00000163508, ENSG00000073861, ENSG00000138755, ENSG00000169245, ENSG00000139187, ENSG00000026508, ENSG00000104894 |
| Th1 signature | IFNG, IL2, CD40LG, IL21, TBX21, STAT4, IL12RB2 | ENSG00000111537, ENSG00000109471, ENSG00000102245, ENSG00000138684, ENSG00000073861, ENSG00000138378, ENSG00000081985 |
| Th2 signature | IL4, IL5, IL13, IL10, GATA3, CCR4 | ENSG00000113520, ENSG00000113525, ENSG00000169194, ENSG00000136634, ENSG00000107485, ENSG00000183813 |
| Treg | FOXP3, CTLA4, IL10, TNFRSF18, CCR8, IKZF4, IKZF2 | ENSG00000049768, ENSG00000163599, ENSG00000136634, ENSG00000186891, ENSG00000179934, ENSG00000123411, ENSG00000030419 |
| Treg and Th2 traffic | CCL17, CCL22, CCL1, CCL28, CCR4, CCR8, CCR10 | ENSG00000102970, ENSG00000102962, ENSG00000108702, ENSG00000151882, ENSG00000183813, ENSG00000179934, ENSG00000184451 |
| Tumor proliferation rate | MKI67, ESCO2, CETN3, CDK2, CCND1, CCNE1, AURKA, AURKB, E2F1, MYBL2, BUB1, PLK1, CCNB1, MCM2, MCM6 | ENSG00000148773, ENSG00000171320, ENSG00000153140, ENSG00000123374, ENSG00000110092, ENSG00000105173, ENSG00000087586, ENSG00000178999, ENSG00000101412, ENSG00000101057, ENSG00000169679, ENSG00000166851, ENSG00000134057, ENSG00000073111, ENSG00000076003 |
| Tumor-associated Macrophages | IL10, MRC1, MSR1, CD163, CSF1R, IL4I1, SIGLEC1, CD68 | ENSG00000136634, ENSG00000260314, ENSG00000038945, ENSG00000177575, ENSG00000182578, ENSG00000104951, ENSG00000088827, ENSG00000129226 |
